# Predicting High Excess Risk of Hyponatremia Among Thiazide Users

**DOI:** 10.1101/2024.10.06.24314957

**Authors:** Niklas Worm Andersson, Kim Daniel Jakobsen, Anders Hviid, Bjarke Feenstra, Mads Melbye, Jan Wohlfahrt, Marie Lund

## Abstract

**Importance:** Hyponatremia is a potential serious adverse drug reaction to treatment with thiazide diuretics. Whether individuals with a high risk of developing thiazide-induced hyponatremia can be predicted before treatment initiation is unknown.

**Objective:** To identify patients with a high risk of thiazide-induced hyponatremia following treatment initiation.

**Design:** Population-based cohort study, using national register data on demography, comorbidities, comedication, and blood test results to predict individual-level risk of thiazide-induced hyponatremia following treatment initiation based on models trained with the causal forest method. Models were validated in a separate cohort and compared by concordance-for-benefit (C-for-benefit) and calibration plots.

**Setting:** Denmark, 1 January 2014 to 31 December 2020.

**Participants:** Patients were ≥40 years old and new users of thiazide or non-thiazide antihypertensive drugs (renin-angiotensin-system inhibitors or calcium channel blockers) and split into a development (n=185,699; from 2014-2018) and a validation cohort (n=75,030; 2019-2020).

**Exposure:** Thiazide or non-thiazide antihypertensive treatment.

**Main outcome and measure:** Predicted excess risk of moderate-to-severe hyponatremia defined as plasma sodium levels <130 mmol/L, measured within the first 120 days of treatment.

**Results:** Individual-level excess risk could be parsimoniously described by a four-covariate model that included information on age and plasma sodium, hemoglobin, and C-reactive protein levels at baseline with good calibration and C-for-benefit (0.66; 95% CI, 0.66-0.67) in the validation cohort. The average 120-day excess risk of hyponatremia among thiazide-treated patients was 1.8% (95% CI, 1.3% to 2.2%), with individual-level heterogeneity ranging from –1.6% to 15.9%. The 10% of thiazide-treated with the highest excess risk of hyponatremia had an average excess risk of 7.4% (95% CI, 4.4% to 10.5%). If this high-risk group were instead assigned a non-thiazide antihypertensive, the excess risk within the remaining thiazide-treated population would be reduced by 0.7% (95% CI, 0.1% to 1.2%), corresponding to a 38% relative reduction.

**Conclusion and relevance:** The population-level burden of thiazide-induced hyponatremia can potentially be markedly reduced by assigning a small group of patients at highest risk, who would otherwise be assigned a thiazide, to an alternative antihypertensive drug. This high-risk group can be identified using a small set of simple baseline information.

## INTRODUCTION

Thiazide diuretics are widely used to treat high blood pressure and are among the first-line pharmacologic drugs for uncomplicated hypertension in the US and European countries.^1,2^ Hyponatremia, overall defined as plasma sodium levels <135 mmol/L,^3^ is a known adverse drug reaction to treatment with thiazide diuretics.^4^ The clinical manifestation of hyponatremia ranges widely from asymptomatic or non-specific symptoms such as dizziness or fatigue to life-threatening conditions such as general seizures and unconsciousness and may be influenced by both the rapidity and degree of sodium depletion.^3,5^

Thiazide diuretics are a well-established cause of drug-induced hyponatremia and have been reported to contribute to 28% of all hospital-associated cases of severe hyponatremia (i.e., plasma sodium ≤125 mmol/L), most often occurring within the first weeks to months of treatment.^4,6–8^ Currently, it is not possible to determine whether a patient is at high risk of thiazide-induced hyponatremia before treatment initiation. Such information on potential treatment harm, however, could support the initial choice of antihypertensive drug and/or monitoring choices.

Estimates of the risk of thiazide-induced hyponatremia in different patient populations vary significantly. Notably, studies have reported an incidence of 30% among outpatient thiazide users^9^ and 4.1% among trial participants^10^. This suggests that the effect of thiazides on hyponatremia risk varies by patient characteristics, that is, heterogeneity of treatment effect. Notably, such reported population averages of treatment effects most likely do not represent the treatment effect in individual patients.^11,12^ Additionally, the traditional methods for identifying high-risk individuals stratify by one factor at a time, which does not allow for the possible complex interplay between multiple factors contributing to treatment harm at the individual patient level.^11,13^ Using causal machine-learning methods enables moving from such treatment effect averages across populations or broad subgroups towards individual-level predictions of effects of a given treatment.^13–15^ This could be leveraged to identify patients at high risk of treatment harm before initiating treatment. Consequently, this Danish nationwide cohort study sought to identify patients with high excess risk of moderate-to-severe hyponatremia (plasma sodium <130 mmol/L) within the first 120 days of thiazide treatment based on individual-level patient characteristics.

## METHODS

### Data Sources and Study Population

This nationwide cohort study used data from all individuals in Denmark aged ≥40 years from 1 January 2014 to 31 December 2020 identified using the Danish Civil Registration System^16^. The unique personal identifier from this register allowed linking of individual-level information on antihypertensive and other prescription drug use, comorbidities, socio-economic characteristics, and laboratory blood test results from other nationwide registers of healthcare and demography.^17–23^

Patients initiating thiazide use (bendroflumethiazide or hydrochlorothiazide—hydrochlorothiazide as a combination pill with a renin-angiotensin-system inhibitor) were compared with patients initiating non-thiazide antihypertensive drug use (renin-angiotensin-system inhibitor or calcium channel blocker [i.e., dihydropyridines]) for hypertension. Using the Register of Medicinal Product Statistics^18^, treatment assignment to new use of thiazide or non-thiazide antihypertensive drugs was designated by the first filled prescription of a study drug within the study period, and this date served as the index date (Table S1). Eligibility criteria (Table S1) and confounders and/or candidate predictors of treatment harm (Table S2) were assessed at the index date; the latter comprised a broad list of prespecified variables (n=66) covering demography and medical history, including comorbidities, comedications, and laboratory blood test results.

Each eligible patient was followed per-protocol from the index date for 120 days for an outcome event of moderate-to-severe hyponatremia, defined as a plasma sodium level measurement <130 mmol/L, ascertained via the Register of Laboratory Results for Research^23^ (Table S1). Accordingly, patients were censored if they experienced an outcome event, deviated from assigned treatment (by stopping assigned use or starting use of any other antihypertensive drug), died, disappeared/emigrated, moved to the Central Denmark Region (where data on laboratory results for the study period were incomplete), reached day 120 of follow-up, or if the study period ended, whichever occurred first.

The study was approved by Statens Serum Institut’s Data Protection and Information Security Department. Under Danish law, neither informed consent nor scientific ethics committee approval is required for strictly register-based studies. Data cells with less than five subjects (but not zero) were not reported to ensure privacy preservation.

### Statistical Analysis

#### Study Design

The causal forest method^14,24^ was used to estimate the individual-level 120-day excess risk of hyponatremia in thiazide users (also termed thiazide-induced hyponatremia). Data was split into a development (years 2014-2018) and a validation (years 2019-2020) cohort to investigate the temporal generalizability of the models to future prediction scenarios. Multiple imputations accounted for missing data under the assumption of missingness at random (given covariates), and results were pooled from each imputed dataset (see Supplementary Methods for further details).

#### Model specification

The utilized causal forest method is an implementation of generalized random forest^14,24^ that employs the R-Learner^25^, building upon Robinson’s transformation^26^, to define a local estimating equation to estimate excess risk as a function of baseline covariates. A random forest is a tree-based, non-parametric algorithm that builds an ensemble of decorrelated decision trees by repeatedly selecting a random subset of the sample for each tree and randomly selecting variables for each split within each tree. The causal forest method requires propensity score and mean outcome predictions as input. These were obtained from regression forests trained on all 66 covariates.

#### Model Development

Pilot analyses using five multiple imputations were conducted to initially evaluate the discrimination and calibration of a wide range of models differing in their covariate compositions. From a model trained on all 66 covariates, measures of variable importance and rank-weighted average treatment effect were used to prioritize covariates for inclusion in the different models.^27^ A simple weighted average of split frequencies at each depth of the trees was used to quantify the variable importance. This is based on the observation that splits in causal trees are chosen to maximize heterogeneity in the excess risk. During this phase, models with few variables performed similarly to the full 66-covariate model and were considered to have higher potential for implementation in clinical practice due to their simplicity. Consequently, models with 1, 2, 3, 4, and 5 covariates, prioritized by variable importance, were selected for final model training with 30 multiple imputations and subsequent validation. These selected parsimonious models were supplemented with a 20-covariate model and the full 66-covariate model to ensure generalizability (i.e., seven selected models were carried forward for model validation in the validation cohort). See Supplementary Methods for additional details.

#### Model Validation

Model performance was evaluated for all seven selected and developed models considering discrimination, calibration, and estimated population benefit. In the development cohort, performance was initially assessed using out-of-bag prediction of the risk of thiazide-induced hyponatremia. Discrimination was assessed by model-based concordance-for-benefit (C-for-benefit)^28^. Calibration was assessed using a plot of the observed excess risk against the predicted excess risk and obtaining the intercept and slope parameters of a fitted linear regression (see Supplementary Methods for details).^29^ Population benefit was assessed by estimating the benefit of switching the treatment assignment to a non-thiazide antihypertensive drug for patient groups with a high risk of thiazide-induced hyponatremia. Under this treatment assignment condition, (a) the population excess risk reduction among thiazide-treated and (b) the number-needed-to-not-treat with a thiazide to prevent one thiazide-induced hyponatremia event were estimated. Motivated by the excess risk distribution in the respective cohort, the 90th and 95th percentiles were used to define these high-risk patient groups for model comparisons.

Agreement between the selected models with respect to identifying these high-risk (and non-high risk) patients was assessed by alternately treating each model as the gold standard to calculate the proportion of agreement between the high-risk group of each other model and the high-risk group of the gold standard model. Correlations between the predicted individual-level excess risks from each pair of models were also reported.

#### Prediction of Individual-Level Excess Risk and Average Excess Risk in Subgroups

One of the seven models was selected for further characterization based on calibration and discrimination performance. For this model, the distribution of the predicted individual-level excess risk was presented together with the average excess risks in subgroups defined by each model variable separately and all two-way combinations. Additionally, the potential average excess risk reduction in the thiazide-treated population, achievable by gradually assigning a non-thiazide drug to patients with the highest excess risks of hyponatremia, was calculated.

#### Additional Analyses

Sensitivity analyses included comparing the performance with models developed for the two thiazide subtypes separately and using a complete case cohort (since multiple imputations potentially could cause poorer model prediction when validated). Lastly, the predicted average excess risk was assessed for categories of *a priori* selected variables, combining the development and validation cohorts.

This study is reported according to the recommendations of TRIPOD+AI (Transparent Reporting of a multivariable prediction model for Individual Prognosis Or Diagnosis).^30^

## RESULTS

### Cohort Characteristics

The study cohort totaled 260,729 new users of thiazide or non-thiazide drugs and was split into a development (n=185,699; 31,480 thiazide vs 154,219 non-thiazide users) and a validation cohort (n=75,030; 6,220 thiazide vs 68,810 non-thiazide users). Baseline characteristics are presented in Table 1 (see Figure S1 for cohort construction), with missing data summarized in Table S2. During the 120-day follow-up period since treatment start, 1362 and 501 patients in the development- and validation cohort, respectively, developed hyponatremia (sodium <130 mmol/L).

**Table 1.**
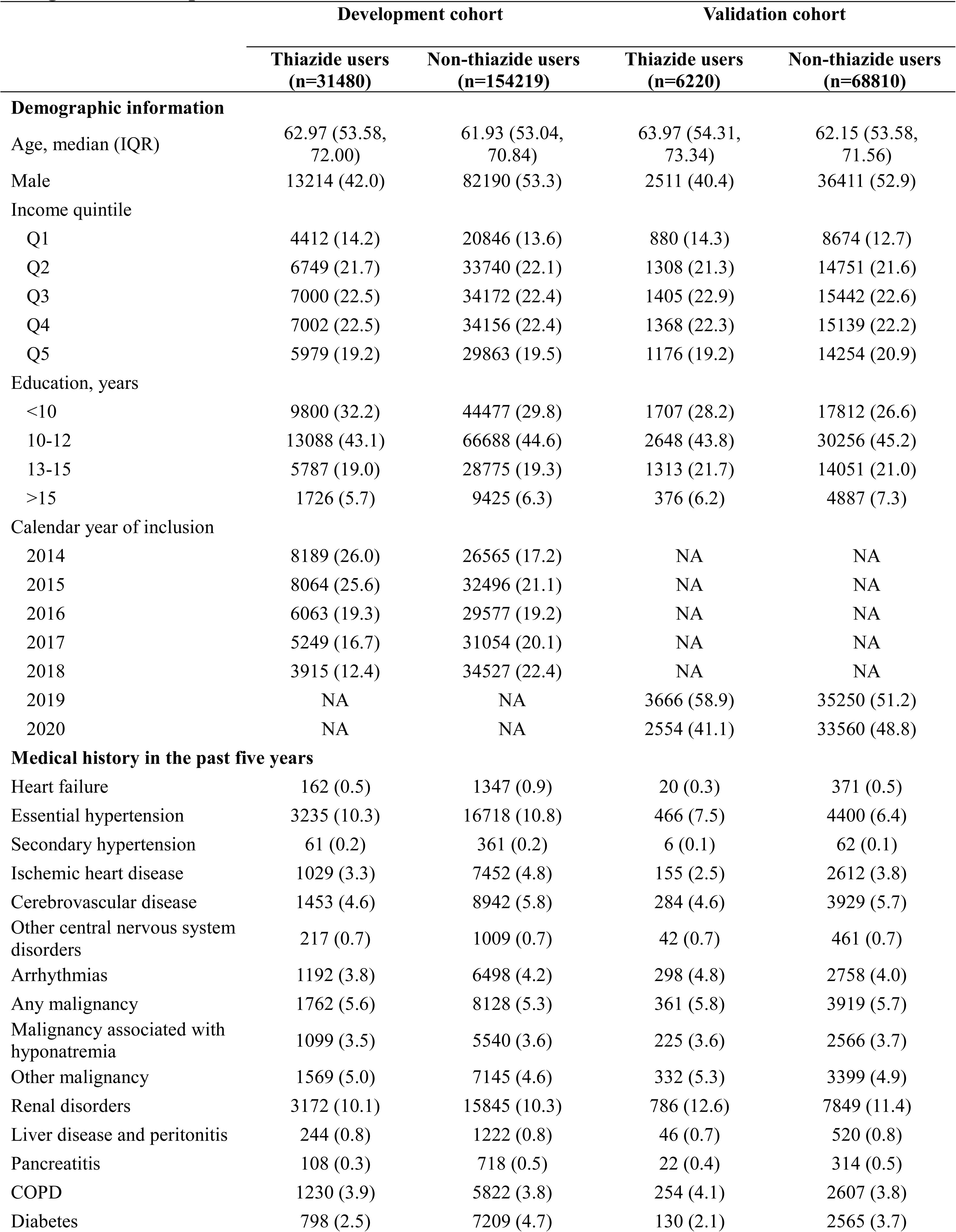
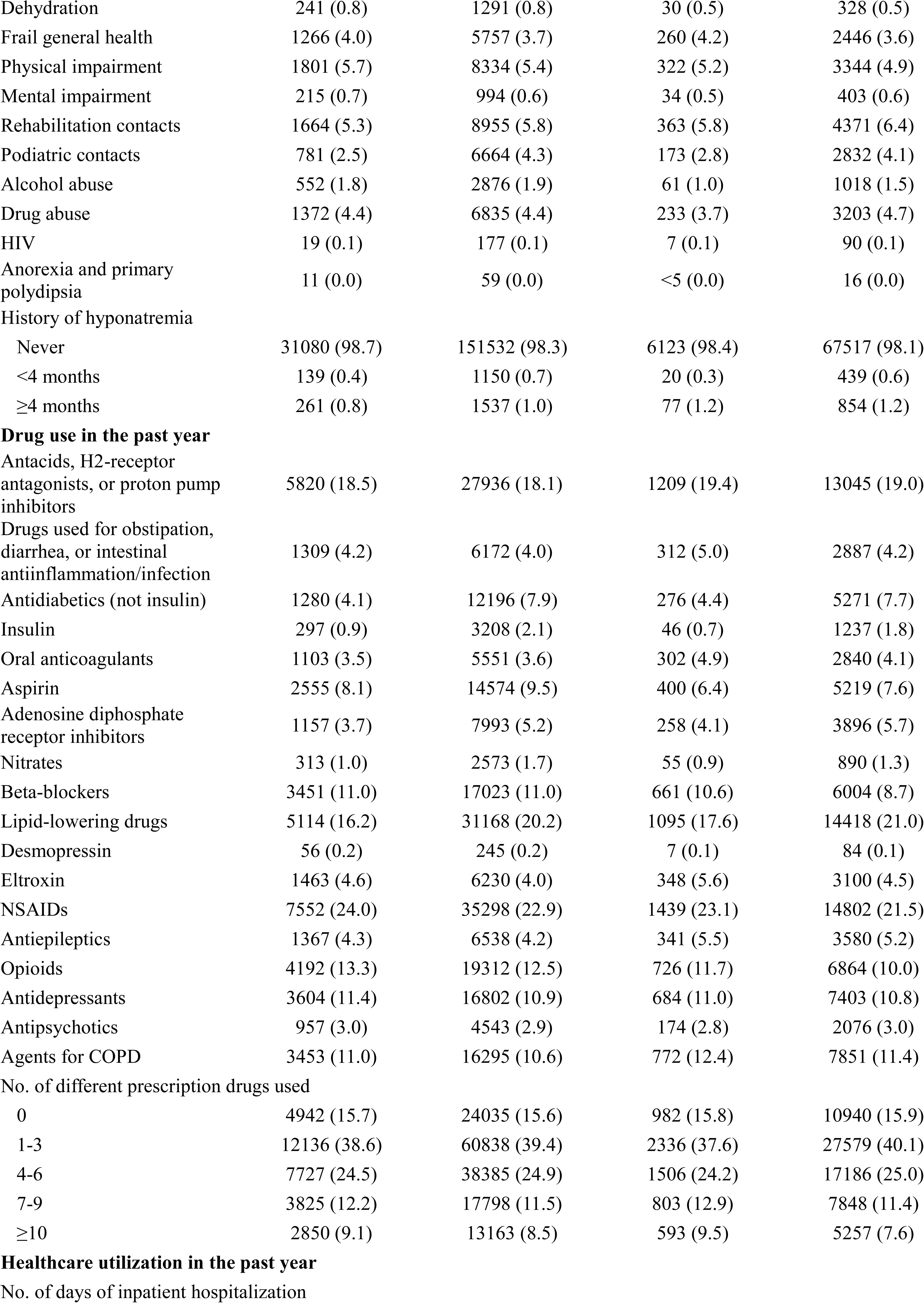
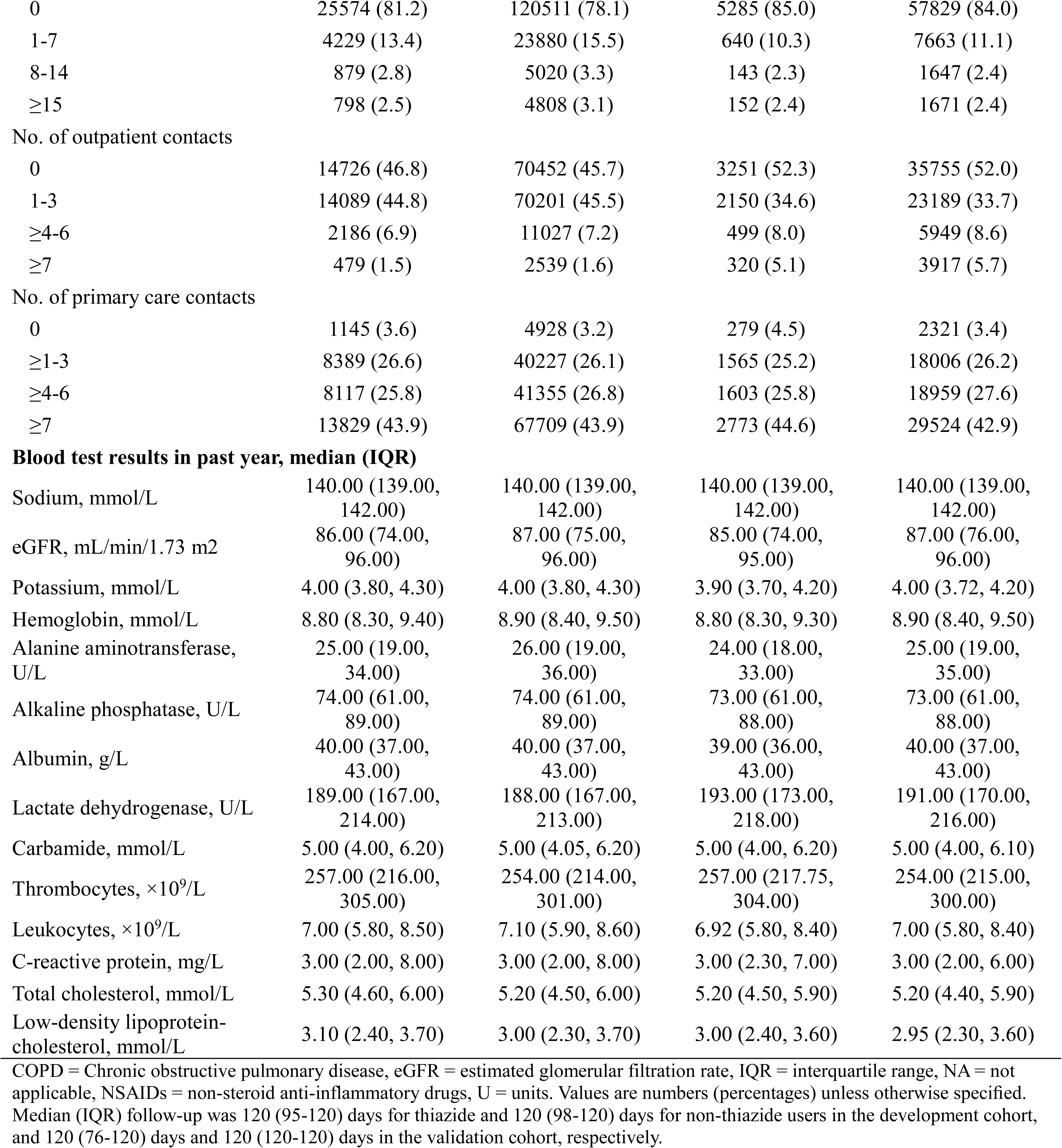
Baseline characteristics of new users of thiazide and non-thiazide antihypertensive drug in the development and validation cohort.

### Discrimination and calibration

Seven different causal forest models were trained in the development cohort (Table S3 lists variables prioritized in the various models and their relative importance); discrimination, calibration, and population benefit measures are presented in Tables S4-S5 and Figure S2.

In the validation cohort, all models, except for the one-covariate model, had similar predictive performance with only minor differences (Table 2, Figures S3-S5, and Table S6). Models with many covariates tended to have higher C-for-benefit (ranging from 0.61 to 0.68) but predicted lower excess risks than observed (e.g., the full model had an observed-by-predicted-excess-risk-regression slope coefficient of 2.84, 95% CI 2.14 to 3.54 together with an intercept of -0.01, 95% CI -0.02 to -0.00).

**Table 2.** Discrimination and calibration measures for the seven selected models in the validation cohort.

| **Models** | **C-for-benefit (95% CI)** | **Calibration regression slope-coefficient (95% CI)** | **Calibration regression slope-intercept (95% CI)** |
| --- | --- | --- | --- |
| 1-cov | 0.533 (0.529 to 0.536) | 0.797 (0.228 to 1.366) | 0.017 (0.004 to 0.030) |
| 2-cov | 0.605 (0.599 to 0.612) | 1.244 (0.786 to 1.703) | 0.015 (0.008 to 0.023) |
| 3-cov | 0.659 (0.652 to 0.667) | 1.909 (1.493 to 2.325) | 0.003 (-0.004 to 0.009) |
| 4-cov | 0.664 (0.655 to 0.672) | 1.977 (1.501 to 2.453) | -0.002 (-0.010 to 0.006) |
| 5-cov | 0.674 (0.665 to 0.683) | 2.489 (1.910 to 3.068) | -0.006 (-0.015 to 0.002) |
| 20-cov | 0.684 (0.677 to 0.691) | 3.130 (1.848 to 4.411) | -0.013 (-0.031 to 0.004) |
| 66-cov (full) | 0.666 (0.661 to 0.670) | 2.837 (2.138 to 3.535) | -0.012 (-0.023 to -0.001) |

CI = confidence intervals, C-for-benefit = concordance-for-benefit. Models are abbreviated 1-cov to 66-cov according to the number of covariates included in the respective model. The corresponding calibration plots for the slope coefficients and intercepts are shown in Figure S3.

All models had a high degree of agreement in assigning patients to the high-risk groups, and overall correlation in individual-level predicted excess risks (Figures S6-S7 and Table S7).

The parsimonious four-covariate model, which only utilized information about age, plasma sodium, hemoglobin, and C-reactive protein at baseline, appeared to have the best calibration by visual inspection and preserved good discrimination performance in the validation cohort and was therefore selected for further characterization.

### High-risk groups and population benefit

Figure 1 shows the predicted risk of thiazide-induced hyponatremia by the four-covariate model in subgroups defined by combinations of the four utilized baseline variables (Figures S8-S10 present the distributions of the individual-level excess risks by the four variables and in combinations). Overall, the average excess risk was higher for lower baseline sodium and hemoglobin levels and for higher baseline C-reactive protein levels and age. For example, the average predicted excess risk of hyponatremia was >5% when baseline plasma sodium levels were <138 mmol/L and age was ≥70 years.

**Figure 1.**
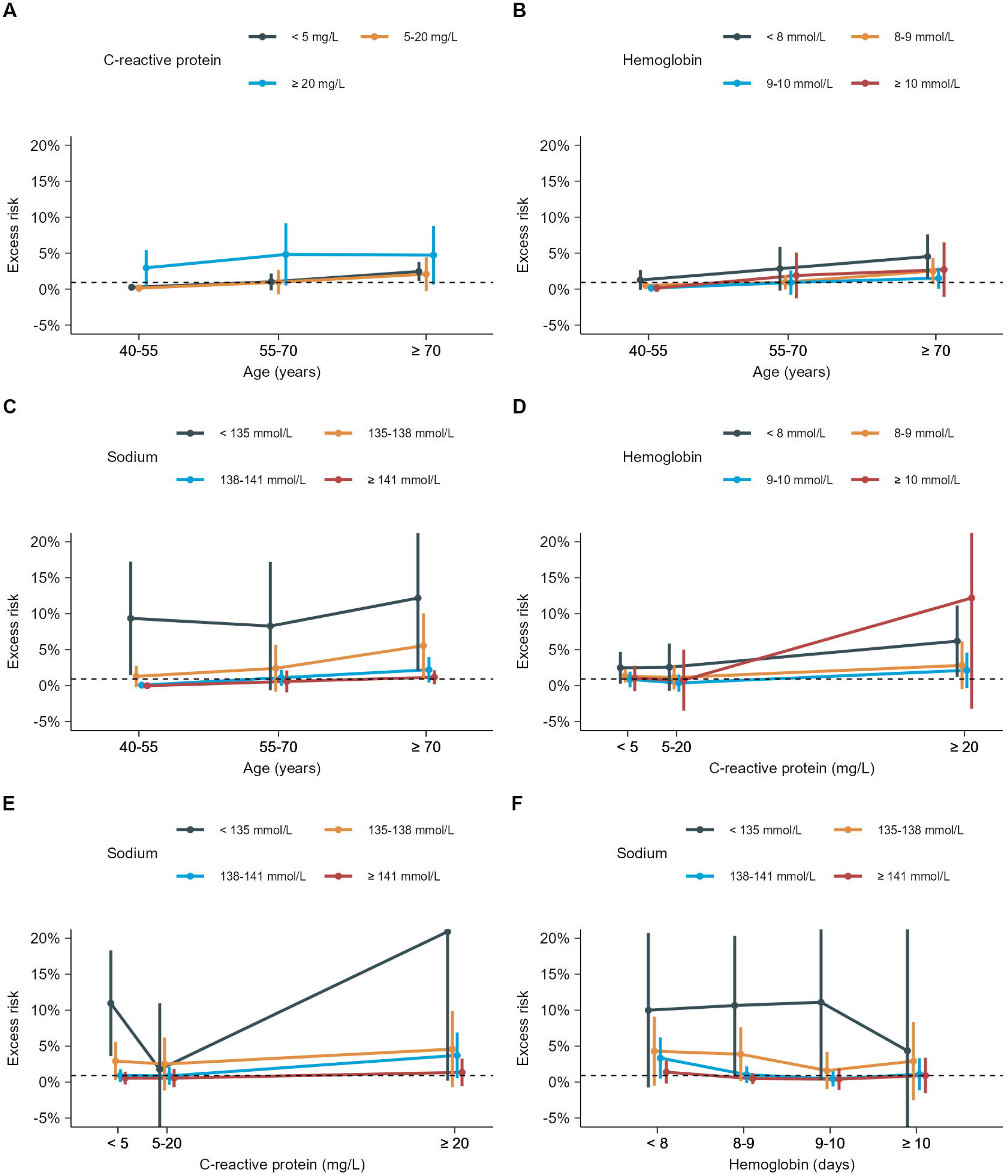
Average excess risk of hyponatremia with thiazide use by combinations of baseline variables. Figure presents the average excess risk and 95% confidence intervals of hyponatremia with thiazide use in the validation cohort for subgroups defined by combinations of baseline plasma sodium, hemoglobin, and C-reactive protein levels and age predicted by the selected four-covariate model (the different two-way combinations are presented in A to F). The distributions of the individual-level excess risk within the covariate-defined subgroups are presented in Figure S8-S10.

The four-covariate model predicted an average risk of thiazide-induced hyponatremia in the thiazide-treated population of 1.8% (95% CI, 1.3% to 2.2%) (Table S8), with individual-level heterogeneity ranging from - 1.6% to 15.9% (Figure 2). The average excess risk for thiazide users with the 10% highest excess risk was 7.4% (4.4% to 10.5%). Assigning non-thiazide treatments to this patient group was estimated to reduce the average excess risk in the thiazide-treated population by 0.7% (95% CI, 0.1% to 1.2%) (Figure 3 and Table S8). This corresponds to a relative reduction in thiazide-induced hyponatremia cases within the entire thiazide-treated population of 38.3%, while the number-needed-to-not-treat with thiazide to prevent one thiazide-induced hyponatremia event would be reduced from 56 (95% CI, 74 to 45) to 13 (95% CI, 23 to 10) patients. Similarly, defining high-risk as the 5% highest excess risk would reduce population burden by 28.1%.

**Figure 2.**
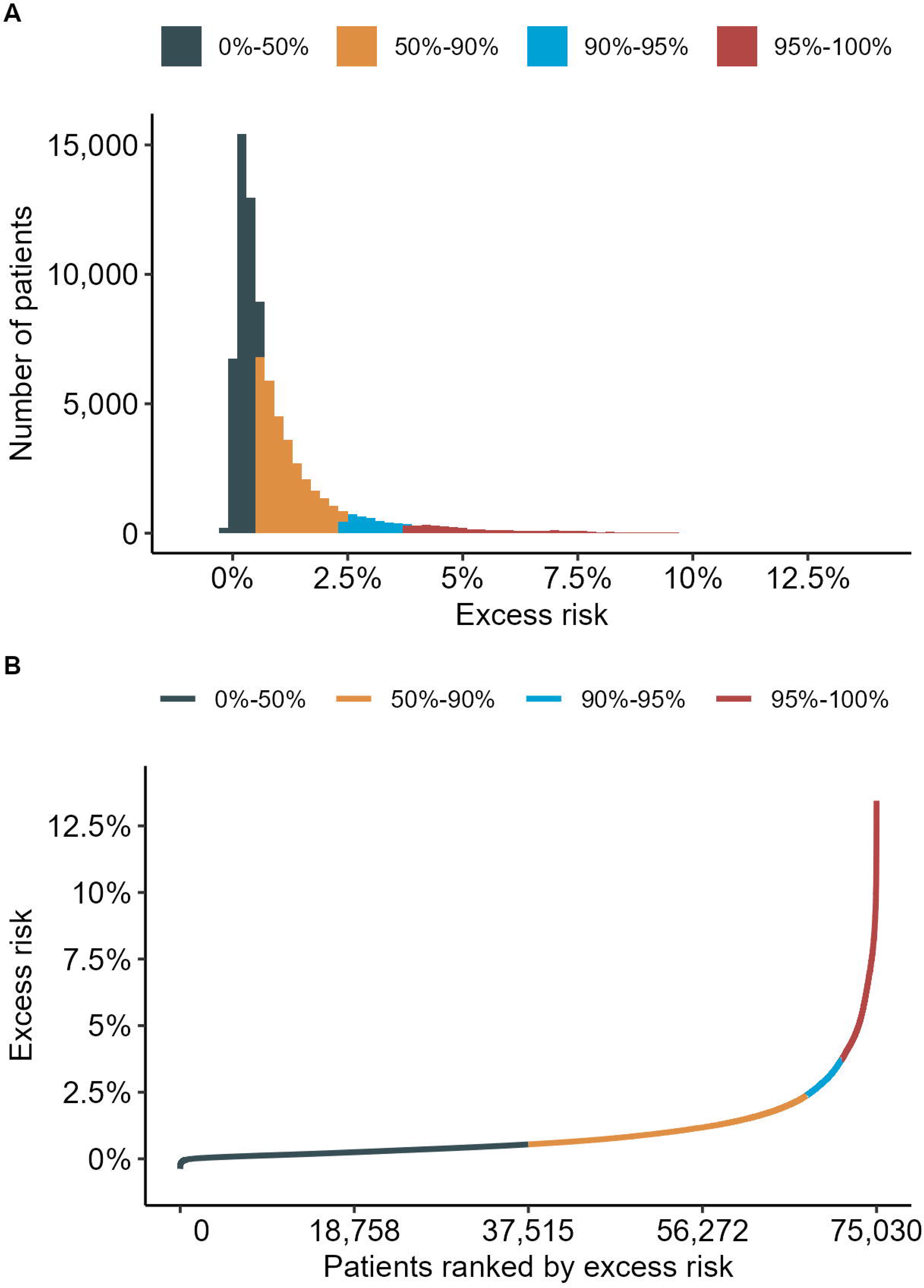
Distribution of individual-level excess risk of hyponatremia with thiazide use. Prediction is performed in the validation cohort using the selected four-covariate model. Figures present: (A) the distribution of the individual-level excess risk of hyponatremia with thiazide use (bin width: 0.2%) and (B) the ranked distribution of the individual-level excess risk of hyponatremia with thiazide use (ascending order).

**Figure 3.**
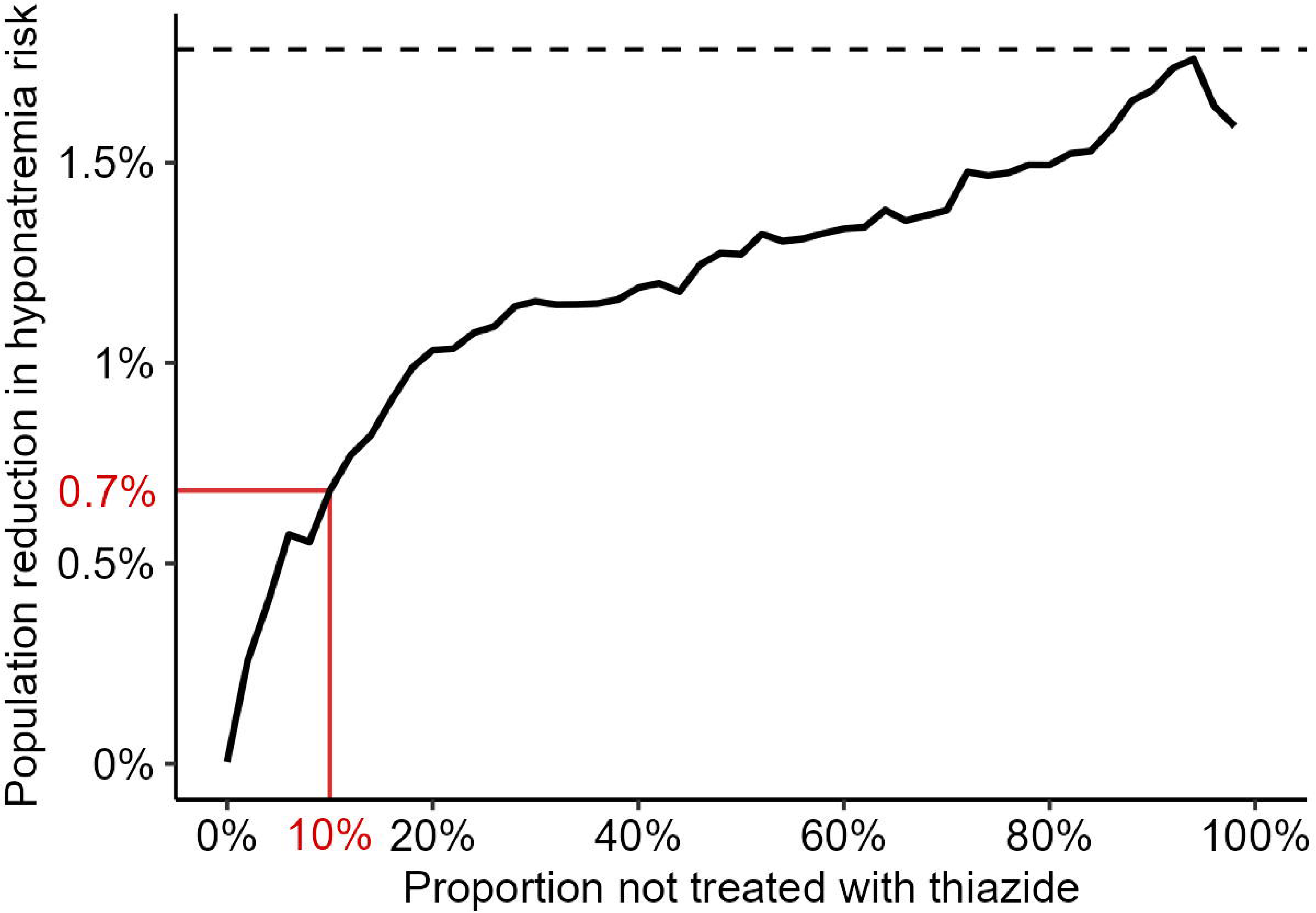
Reduction in population-level risk of thiazide-induced hyponatremia when switching from thiazide to non-thiazide treatment in targeted subpopulations. Figure presents the population reduction in the risk of thiazide-induced hyponatremia as the proportion of patients not treated with thiazides increases, according to their ranked excess risk (descending order). The x-axis reflects the cumulative percentage of individuals excluded from thiazide treatment (i.e., assigned a non-thiazide antihypertensive instead). Population risk reduction was calculated as the decrease in overall risk of thiazide-induced hyponatremia among thiazide treated as higher-risk individuals are gradually excluded from thiazide treatment. The individual-level risk predictions are based on the selected four-covariate model within the validation cohort. Population estimates are calculated by average of individual-level estimates using augmented inverse propensity weighting, which produces asymptotically unbiased, double robust estimates. Consequently, the curve may not be smooth throughout due to the variability introduced by this method, particularly when differences are small. The population reduction in the risk of thiazide-induced hyponatremia when not treating the 10% highest ranked in excess risk is highlighted in red. The population average risk of thiazide-induced hyponatremia (of 1.8%) is shown with a horizontal dashed line.

### Additional Analyses

Compared to the selected final four-covariate model, the prediction performance of models derived for each thiazide subtype separately was similar in the development and for the bendroflumethiazide model in the validation cohort, but the hydrochlorothiazide model was limited by a small treated group in the validation cohort, making it difficult to evaluate the performance of this model (Tables S9-S10 and Figure S11). Findings were similar for models developed using complete cases instead of multiple imputations (Tables S11-S12). Lastly, the average excess risk of hyponatremia with thiazide use was reported across stratified subgroups of 20 *a priori* chosen variables (Table S13). Prior history of dehydration (4.6% [1.6% to 7.5%]) and recent hyponatremia (8.2% [2.1% to 14.2%]) were among those predictors with the highest effect on excess risk of hyponatremia upon thiazide initiation.

## DISCUSSION

Using causal machine learning, this Danish nationwide cohort study predicts that hyponatremia following thiazide initiation can be markedly reduced at the population level by assigning an alternative antihypertensive drug to a group of patients with a high predicted risk of thiazide-induced hyponatremia. Specifically, by use of a parsimonious model, the identified 10% of thiazide users with the highest excess risk of hyponatremia had a predicted average of 7.4%, while the population average was 1.8%. By switching from thiazide to non-thiazide treatment assignment for this high-risk group, the population burden of thiazide-induced hyponatremia would be reduced by 38%.

Baseline plasma sodium level, followed by age, were the two most important variables for determining the heterogeneity in treatment-related harm. Not surprisingly, a higher risk of thiazide-induced hyponatremia was observed with lower baseline sodium and older age. The other two variables in the characterized four-covariate model—hemoglobin and C-reactive protein levels—also predicted higher excess risk at lower hemoglobin and higher C-reactive protein levels. Consistent with prior studies^31–35^, other factors, such as comorbidities, were found to independently influence the risk (e.g., higher excess risk was observed in patients with a history of dehydration or malignancy in conventional subgroup analysis considering one variable at a time). However, adding these other factors to models of the individual risk did not meaningfully improve performance or alter the predicted population benefit. To our knowledge, this is the first study to predict the risk of thiazide-induced hyponatremia in populations and demonstrate the potential for markedly reducing population burden by leveraging simple and accessible baseline information.

Such a parsimonious model, requiring minimal information input, holds realistic promise for adaptation in clinical practice to assist in treatment decision-making and/or to optimize monitoring schedules for adverse effects. Models with fewer covariates (i.e., the two- and three-covariate models) demonstrated only slightly lower discrimination and calibration, offering similar population benefits and may be even better candidates for clinical implementation. This study used the excess risk distribution to define high-risk groups, but other high-risk definitions, such as absolute thresholds, could be easily adapted and relevant depending on the clinical scenario. As many factors may influence antihypertensive treatment choices (e.g., the totality of contraindications, comorbidities, and expected effect and safety), identifying a small group of patients at particularly high risk of this adverse drug reaction appears valuable in aiding clinical care.

This study has limitations. First, this study takes advantage of the fact that information on blood test measurements, medication use, and diagnosed conditions from routine clinical practice is recorded in the national healthcare registers. Using nationwide registers with individual-level data increases the generalizability and mitigates risks of selection bias, information bias, and loss to follow-up. However, the utilized data lack true randomization of treatment assignment. Instead, exchangeability was conditioned on 66 covariates that have potential causal links to both treatment assignment and the risk of hyponatremia. Yet, unmeasured confounding factors, not indirectly accounted for, cannot be excluded, which, if present, will have induced bias in the reported risks. Similarly, unmeasured effect modifiers may exist, potentially implying undetected effect heterogeneity not captured by the included variables. Second, since the data derive from everyday clinical practice, not all blood values were measured for all patients at baseline. However, less than 8% of patients in the validation cohort had missing baseline sodium measurements, and missingness was addressed using multiple imputations. Missing blood values likely indicates that the unobserved values are within the normal range. Still, it is reassuring that comparable results were observed for a model based on complete cases. Third, use of a temporal split-sample design allows for more exploratory and adaptive model development, and it involves two distinct cohorts that can differ more than would be expected by chance alone. This also promotes the likelihood that any inherent unobserved bias will lead to poorer model performance when applied to the validation cohort; however, using data from different sources or geographically distinct cohorts could further empower the robustness of the models. Nonetheless, this study demonstrates that causal forest machine-learning methods can be effectively combined with nationwide observational healthcare data to inform and reduce treatment-related harm in populations. Fourth, this study considers excess risk as the most clinically meaningful measure of treatment harm in this context, and using causal forest models enabled the estimation of this; however, it also challenges model evaluation. To assess calibration, observed counterfactual outcomes were generated by matching thiazide and non-thiazide users, but the quality of this operation is difficult to evaluate since direct individual-level excess risks are unobservable. Fifth, this study exploits the fact that all results from blood tests are registered nationally. However, this makes differences in outcome ascertainment a potential concern. In Denmark, patients treated for hypertension with antihypertensive drugs are recommended to have their electrolyte levels routinely checked, and all healthcare services are free of charge.^19^ Additionally, we recently found no differences in plasma sodium ascertainment by the exposure using the same data.^4^ Potential differences are further mitigated by the active comparator design, inclusion of a wide range of covariates likely also predicting the ascertainment of non-symptomatic plasma sodium, and the restriction to moderate-to-severe hyponatremia (<130 mmol/L) to increase the likelihood of symptomatic manifestations leading to testing and outcome detection.

## CONCLUSION

This Danish nationwide cohort study finds that by assigning a relatively small group of patients at high risk of thiazide-induced hyponatremia to an alternative non-thiazide antihypertensive treatment, the burden of this adverse drug effect can be markedly reduced in populations. These high-risk patient groups can be identified using simple baseline information within a parsimonious causal machine-learning prediction model.

## Author Contributions

Dr Andersson and Mr Jakobsen had full access to all the data in the study and take responsibility for the integrity of the data and the accuracy of the data analysis.

*Concept and design:* All authors.

*Acquisition, analysis, or interpretation of data:* All authors.

*Drafting of the manuscript:* Andersson.

*Critical revision of the manuscript for important intellectual content:* All authors.

*Statistical analysis:* Jakobsen.

*Supervision:* Hviid, Wohlfahrt, Lund.

## Conflicts of Interest and Financial Disclosures

The authors declare no competing interests.

## Funding/Support and Role of Funder/Sponsor

This study was supported by a Data Science Investigator grant from the Novo Nordisk Foundation and grants from Independent Research Fund Denmark, Helsefonden, Dagmar Marshalls Fond, Gangstedfonden, A.P. Møller and Chastine Mc-Kinney Møller Foundation, Brødrene Hartmanns Fond, and Snedkermester Sophus Jacobsen og hustru Astrid Jacobsens Fond. No funder had any role in the design and conduct of the study; collection, management, analysis, and interpretation of the data; preparation, review, or approval of the manuscript; and decision to submit the manuscript for publication.

## Data availability

There are no additional data available. Owing to data privacy regulations in Denmark, the raw data cannot be shared. However, the data are available for research upon reasonable request to the Danish Health Data Authority and within the framework of the Danish data protection legislation and any required permissions from relevant authorities.

## Notes

### Competing Interest Statement

The authors have declared no competing interest.

### Author Declarations

The study was approved by the Data Protection and Information Security Department of Statens Serum Institut. Under Danish law, neither informed consent nor scientific ethics committee approval is required for strictly register-based studies. Data cells with less than five subjects (but not zero) were not reported to ensure privacy preservation.

