## Supplementary material for "Predicting High Excess Risk of Hyponatremia Among Thiazide Users": Table S1

**Supplementary Table S1. Eligibility criteria, exposure, and outcome variable definitions**

| Variable | Data source(s)[1–5] and details[a] | Codes |
|---|---|---|
| **Eligibility criteria** | | |
| Aged 40 years or older | The Civil Registration System. Defined as the index date minus birthdate ≥40 years. | n/a |
| No use of any antihyper-tensive drug use | Register of Medicinal Products Statistics. Defined as no filled prescription for an antihypertensive drug in the past year before the index date. | ATC: C02, C03, C04, C08, C09 |
| No initiation of multiple antihypertensive drugs | Register of Medicinal Products Statistics. Defined as no filling of multiple prescriptions of the studied drugs (i.e., exposure or comparators) on the index date (not ATC: C09BA or C09DA alone but, e.g., if combined with a CCB or a thiazide). | ATC: C03AB01, C09BA, C09DA, C09A, C09C, C08CA |
| Hypertension as the recorded treatment indication | Register of Medicinal Products Statistics. Defined as the recorded indication for the filled prescription of a study drug on the index date. | indo: 0000054, 0000057 |
| Known address during last two years | The Civil Registration System. Defined as registered address in the two years before the index date. | n/a |
| Not living in Central Denmark Region | The Civil Registration System. Defined as the last recorded registered address not in Central Denmark Region before the index date. Because laboratory data was not fully available from this region. | Region code number: '82' |
| No end-stage illness | The National Patient Register. Defined as no diagnosis of any type recorded during any hospital contact in the last five years before the index date. | ICD-10: E41-E43, F051, I702A, R34, R402, R403, R64, R029, Z49, Z991 |
| No adrenal insufficiency disorders (incl. Addison's disease) | The National Patient Register. Defined as no diagnosis of any type recorded during any hospital contact in the last five years before the index date. | ICD-10: E27 |
| No kidney artery stenosis | The National Patient Register. Defined as no diagnosis of any type recorded during any hospital contact in the last five years before the index date. | ICD-10: Q271, I701, N280 |
| No severe kidney disease (i.e., stage 4 or lower) | The National Patient Register and the Register of Laboratory Results for Research. Defined as no diagnosis of any type recorded during any hospital contact or measured estimated glomerular filtration rate <30 ml/min/1.73 $m^2$ in the last five years before the index date. | ICD-10: N184, N185, Z992<br>SKS: BJFD, BJFZ<br>NPU: DNK35131, DNK35302 |
| **Exposure definition** | | |
| Thiazide-user | Register of Medicinal Products Statistics. Defined as the filled prescription on the index date. Thiazide drugs consisted of bendroflumethiazide and hydrochlorothiazide, the latter in form of a combination pill also containing a renin-angiotensin system inhibitor (hydrochlorothiazide in single-agent tablet has seen only limited use in Denmark during the study period). | ATC: C03AB01, C09BA, C09DA |
| Non-thiazide user | Register of Medicinal Products Statistics. Defined as the filled prescription on the index date. Non-thiazide drugs consisted of renin-angiotensin system inhibitors (comprising angiotensin-converting enzyme inhibitors and angiotensin receptor blockers) and calcium-channel blockers (i.e., dihydropyridines). | ATC: C09A, C09C, C08CA |
| **Outcome definition** | | |
| Hyponatremia | Register of Laboratory Results for Research. Defined as laboratory blood test results with plasma sodium | NPU: NPU03429 |

| | |
|---|---|
| | <130 mmol/L within the first 120 days from the index date. |

ATC = anatomic therapeutic chemical classification, ICD-10 = International Classification of Diseases System, version 10, indo = indication code, n/a = not applicable, NPU = Nomenclature for Properties and Units, and SKS = Danish abbreviation for the Danish Medical Classification System.
[a]The index date was defined as the date of the first filled prescription of a study drug during the study period. Any diagnosis denotes A- and B- (primary and secondary) diagnoses. Any hospital contact denotes inpatient, outpatient, and emergency department contacts.

### Supplementary Table S2. Covariate definitions

| Variable | Values | Codes | % missing (development; validation cohort) |
|---|---|---|---|
| **Demographic information[a]** | | | |
| Age | Continuous | n/a | 0; 0 |
| Sex | Binary: female/male | n/a | 0; 0 |
| Household income, quintiles | Categorical (5 levels): 1, 2, 3, 4, 5 | n/a | 1.0; 0.8 |
| Education, years | Categorical (4 levels): <10, 10-12, 13-15, >15 | n/a | 3.2; 2.6 |
| Calendar year of inclusion | Categorical (7 levels): 2014, 2015, 2016, 2017, 2018, 2019, 2020 | n/a | 0; 0 |
| **Medical history in the past five years[b]** | | | |
| Heart failure | Binary: yes/no | I110, I130, I132, I420, I426, I427, I428, I429, I500, I501, I509, J819 | 0; 0 |
| Essential hypertension | Binary: yes/no | I10 | 0; 0 |
| Secondary hypertension | Binary: yes/no | I15 | 0; 0 |
| Ischemic heart disease | Binary: yes/no | I20-I25 | 0; 0 |
| Cerebrovascular disease | Binary: yes/no | I61, I63, I64, I676, G45 | 0; 0 |
| Other central nervous system disorders | Binary: yes/no | E23, G35, G610, G91, G935, G936, Q03 | 0; 0 |
| Arrhythmias | Binary: yes/no | I48, I49, Z950 | 0; 0 |
| Any malignancy | Binary: yes/no | C00-C97 (not C44) | 0; 0 |
| Malignancy associated with hyponatremia | Binary: yes/no | C00-C41, C45-C49, C51-C72, C81-C86, C884 | 0; 0 |
| Other malignancy | Binary: yes/no | C50, C73-C80, C88-C97 (not C884) | 0; 0 |
| Renal disorders | Binary: yes/no | I120, I131, N00, N01, N03, N04, N05, N17, N181, N182, N183, N19, N26<br>NPU: DNK35131, DNK35302 (30- <60 ml/min/1.73 $m^2$) | 0; 0 |
| Liver disease and peritonitis | Binary: yes/no | K65, K70-K77, I982, Z944, D684C | 0; 0 |
| Pancreatitis | Binary: yes/no | K85, K860, K861 | 0; 0 |
| Chronic obstructive airway disorder | Binary: yes/no | J44, J45, J46 | 0; 0 |
| Diabetes | Binary: yes/no | E10-E14 | 0; 0 |
| Dehydration | Binary: yes/no | E86 | 0; 0 |
| Frail general health | Binary: yes/no | H54, L89, L97, R32, R53, R54, R630, R633, R636, R634, Z74, Z96 | 0; 0 |
| Physical impairment | Binary: yes/no | R671-R679, R26, M625, Z993, R296, S12, S220, S221, S320, S422-S424, S52, S720-S722, T08 | 0; 0 |
| Mental impairment | Binary: yes/no | F00-F03, G30 | 0; 0 |
| Rehabilitation contacts | Binary: yes/no | Z50 | 0; 0 |
| Podiatric contacts | Binary: yes/no | SPEC2: 54, 59, 60 | 0; 0 |
| Alcohol abuse | Binary: yes/no | F10<br>ATC: N07BB01, N07BB03 | 0; 0 |
| Drug abuse | Binary: yes/no | F11-F16, F18-F19, T40, T43, R781-R785, | 0; 0 |

| Variable | Values | Codes | % missing (development; validation cohort) |
|---|---|---|---|
| | | Z722 | |
| HIV | Binary: yes/no | B20-B24 | 0; 0 |
| Anorexia and primary polydipsia | Binary: yes/no | R630, R631 | 0; 0 |
| History of hyponatremia | Categorical: never, within 4 months, and >4 months before the index date. | E871A, E222A<br>NPU: NPU03429 (<130 mmol/L) | 0; 0 |
| **Drug use in the past year**[c] | | | |
| Antacids, H2-receptor antagonists, or proton pump inhibitors | Binary: yes/no | A02A, A02BA, A02BC | 0; 0 |
| Drugs used for obstipation, diarrhea, or intestinal antiinflammation/infection | Binary: yes/no | A06, A07 | 0; 0 |
| Antidiabetics (not insulin) | Binary: yes/no | A10B | 0; 0 |
| Insulin | Binary: yes/no | A10A | 0; 0 |
| Oral anticoagulants | Binary: yes/no | B01AA, B01AE07, B01AF | 0; 0 |
| Aspirin | Binary: yes/no | B01AC06 | 0; 0 |
| Adenosine diphosphate receptor inhibitors | Binary: yes/no | B01AC04, B01AC22, B01AC24 | 0; 0 |
| Nitrates | Binary: yes/no | C01DA | 0; 0 |
| Beta-blockers | Binary: yes/no | C07 | 0; 0 |
| Lipid-lowering drugs | Binary: yes/no | C10 | 0; 0 |
| Desmopressin | Binary: yes/no | H01BA02 | 0; 0 |
| Eltroxin | Binary: yes/no | H03AA01 | 0; 0 |
| NSAIDs | Binary: yes/no | M01A (not M01AX) | 0; 0 |
| Antiepileptics | Binary: yes/no | N03A | 0; 0 |
| Opioids | Binary: yes/no | N02A | 0; 0 |
| Antidepressants | Binary: yes/no | N06A | 0; 0 |
| Antipsychotics | Binary: yes/no | N05A | 0; 0 |
| Agents for obstructive airway disease | Binary: yes/no | R03 | 0; 0 |
| No. of different prescription drugs used | Categorical: 0, 1-3, 4-6, 7-9, ≥10 | All ATC codes | 0; 0 |
| **Healthcare utilization in the past year(s)**[d] | | | |
| Days of hospitalization in the past 1 year before the index date | Categorical: 0, 1-7, 8-14, ≥15 | 2014-2019: c_pattyp: 0; 2020: ≥12 hours or overnight stay | 0; 0 |
| No. of outpatient contacts in the past 1 year before the index date | Categorical: 0, 1-3, 4-6, ≥7 | 2013; c_pattype: 2<br>2014-2019; c_pattype: 2 and c_indm: 2; 2020: <12 hours and not overnight stay | 0; 0 |
| No. of primary care contacts in the past 2 years before the index date | Categorical: 0, 1-3, 4-6, ≥7 | Kontakt: 1 and ydertype: 01-05 and SPEC6: 70-80xxxx | 0; 0 |
| **Blood test results in the past year**[e] | | | |
| Sodium | Continuous | NPU03429 | 22.0; 7.6 |
| eGFR | Continuous | DNK35131, DNK35302 | 20.0; 6.6 |
| Potassium | Continuous | NPU03230 | 32.7; 8.9 |
| Hemoglobin | Continuous | NPU02319 | 37.2; 15.0 |
| Alanine aminotransferase | Continuous | NPU19651 | 24.0; 8.9 |
| Alkaline phosphatase | Continuous | NPU27783, NPU19655 | 32.4; 19.9 |
| Albumin | Continuous | NPU19673, NPU01132 | 48.8; 32.3 |
| Lactate dehydrogenase | Continuous | NPU19658 | 62.9; 47.2 |

| Variable | Values | Codes | % missing (development; validation cohort) |
|---|---|---|---|
| Carbamide | Continuous | NPU01459 | 74.0; 59.3 |
| Thrombocytes | Continuous | NPU03568 | 38.2; 22.7 |
| Leukocytes | Continuous | NPU02593 | 41.6; 20.4 |
| C-reactive protein | Continuous | NPU01423, NPU19748 | 47.4; 28.5 |
| Cholesterol | Continuous | NPU01566, NPU18412 | 38.8; 15.2 |
| LDL-C | Continuous | NPU01568, NPU10171 | 41.0; 27.8 |

ATC = anatomic therapeutic chemical classification, eGFR = estimated glomerular filtration rate, LDL-C = low-density lipoprotein-cholesterol, n/a = not applicable, NPU = Nomenclature for Properties and Units, NSAIDS = non-steroid anti-inflammatory drugs, SPEC2 = two-digit administrative specialty code, SPEC6 = six-digit administrative specialty code.

The index date was defined as the date of the first filled prescription of a study drug during the study period.

[a]Variables were obtained from the Civil Registration System[1] while household income and education level were obtained from Statistics Denmark[6,7] using the most recently recorded information before the index date.

[b]Variables were obtained from the National Patient Register[2], defined as any recorded A- or B-diagnosis during any hospital contact within the past five years before the index date. The Register of Laboratory Results for Research[3,4] was also used for defining renal disorders (as any blood test results with an eGFR measurement between 30 and <60 ml/min/1.73 $m^2$ in the past five years before the index date) and history of hyponatremia (as any blood test results with a plasma sodium measurement <130 mmol/L in the past five years before the index date). Alcohol abuse was also defined by filled prescriptions within the past year before the index date obtained from the Register of Medicinal Product Statistics[5]. Podiatric contacts were obtained from the Danish National Health Insurance Service Register[8] and defined as any recorded contact five years before the index date. Codes are International Classification of Diseases System, version 10, unless otherwise stated.

[c]Variables were obtained from the Register of Medicinal Product Statistics and defined by filled prescriptions within the past year before the index date. Codes are anatomic therapeutic chemical classification codes.

[d]Hospital contact variables were obtained from the National Patient Register, and primary care variables were obtained from the Danish National Health Insurance Service Register, defined by the number of recorded contacts in the past year(s) before the index date.

[e]Variables were obtained from the Register of Laboratory Results for Research and defined by the last available laboratory blood test results within the past year before the index date. Codes are Nomenclature for Properties and Units codes.

### Supplementary Methods

*Supplementary Model Development*

Before training the causal forest, a survival forest, to account for outcome censoring, and regression forests, to obtain propensity score and mean outcome predictions, were trained on all 66 covariates.

The trained survival forest predicted the survival probabilities of the censoring process. These probabilities were used to define inverse probability of non-censoring weights, where each non-censored case was weighted by the inverse probability of non-censoring at the time of the event. If the event was hyponatremia within the 120 day-period, the event time was the first day of hyponatremia. If the event was no hyponatremia within the 120 day-period, the event time was day 120. The weights were used as sample weights for the regression forests that obtained the 1) propensity of thiazide initiation and 2) propensity of observing hyponatremia during follow-up. These estimates were taken forward to train seven different selected causal forest models in the development cohort.

All forest models (survival, regression, or causal) were trained with clustering on calendar year of drug initiation. This was done because we want results to generalize beyond the calendar years included in this study. Clustering in forest models entails first drawing clusters of subsamples and then drawing at random from each selected cluster. The number of trees grown was 2000. The maximum split imbalance was set to 0.0002, chosen to allow splits on all covariates. The remaining hyperparameters were set to the default values from the *grf* R-package[9] when training survival forests. For the remaining forest models, *grf* 's build-in hyperparameter tuning was used with 100 random draws of possible hyperparameter values, 200 trees in each forest used to compute out-of-bag errors, and 1000 random draws of hyperparameter values to evaluate the predicted smoothed error. If tuning did not improve error over default parameters in the causal forest model trained with all covariates, default values were used for all seven selected causal forest models.

*Supplementary Methods for Multiple Imputation*

To account for missing covariate information, multiple imputation was used. Five imputations were used during the pilot analyses, and 30 imputations were used for the seven selected models trained for subsequent external validation. Missing information was present for income, education, and the blood test results (i.e., sodium, estimated glomerular filtration rate, potassium, hemoglobin, alanine aminotransferase, alkaline phosphatase, albumin, lactate dehydrogenase, carbamide, thrombocytes, leukocytes, C-reactive protein, total cholesterol, and low-density-lipoprotein cholesterol levels). Income and education were included as multilevel factors and were imputed using proportional odds logistic regression models. The remaining covariates were included as continuous variables and were imputed using predictive mean matching with a donor pool of size five. The multiple imputation algorithm was run for 30 iterations. Missing information was imputed separately on the development and validation cohorts. Model training was performed separately in each imputed dataset.

All results were first obtained from each of these models and were then pooled to obtain a final result. To obtain excess risks in the validation cohort, the excess risk within each of the 30 imputed validation cohorts was predicted from the models trained on each of the 30 imputed development cohorts and then pooled. The excess risk estimates on each of the 30 imputed validation cohorts were then pooled to obtain a final prediction.

*Supplementary Methods for Variable Importance*

To quantify the importance of each covariate used in a causal forest model with respect to estimating the excess risk of hyponatremia with thiazide use, the frequency of splits along each covariate was recorded at each depth of the trees. Splits in causal trees are chosen to maximize heterogeneity, so the number of splits along each covariate can be used as an indicator of the influence of each covariate on the excess risk estimates. At each tree depth, the number of potential splits to consider is double the number of potential splits at the previous depth. To account for this, the frequency of splits was normalized to the total number of splits at each depth. Since causal forest greedily selects splits with the largest heterogeneity between the leaf nodes, splits higher up the tree carry more importance to the final predictions. This was incorporated by weighting with the reciprocal squared of the tree depth and only considering the first four layers of the causal trees, in accordance with the *grf* defaults[9]. With these choices, 70% of the weight is put on the first layer of each tree, 18% on the second layer, 8 % on the third layer and 4% on the fourth layer. Choosing a high weight on the first- and second-layers results in a higher separation of the most important variables, making it suitable for variable selection. With the chosen weight decay, including depths beyond depth four has negligible effect on the final importance measure as the weights at further depths become negligible (2.7% at depth five, 1.9% at depth six). The weighted frequencies at each depth of the tree were averaged to obtain the final importance measure.

*Supplementary Methods for Model Calibration*

Calibration was assessed by comparing observed and predicted excess risk.[10] For this purpose, patients were aggregated into 10 groups according to the predicted excess risk of hyponatremia (as well as into 5 and 20 groups in a sensitivity analysis). To validly rank patients from the development cohort, the cohort was split randomly into five folds, predicting excess risk in each fold using a causal forest model trained on the remaining folds. To validly rank patients from the validation cohort, the excess risk of hyponatremia was predicted on each validation sample using the developed models. Predicted excess risk in each aggregated group was obtained using the augmented inverse propensity weighting estimator.[11] The excess risk for each patient is inherently unobservable, as individual patient-level outcomes are only observed for the treatment they received, not the counterfactual treatment. To obtain an estimated counterfactual outcome, each thiazide user was matched 1:1 with a non-thiazide user on background risk of hyponatremia, while each non-thiazide user was matched 1:1 with a thiazide user on exposed risk of hyponatremia.[12] Matching was performed with replacement; matches were chosen among all patients from the opposite treatment group (not only patients in the same ranking group). Calibration was assessed by plotting the aggregated observed excess risk against the predicted excess risk for these 10 groups and by the intercept and slope parameters of a linear regression.

### Supplementary Figure S1. Cohort construction flow chart

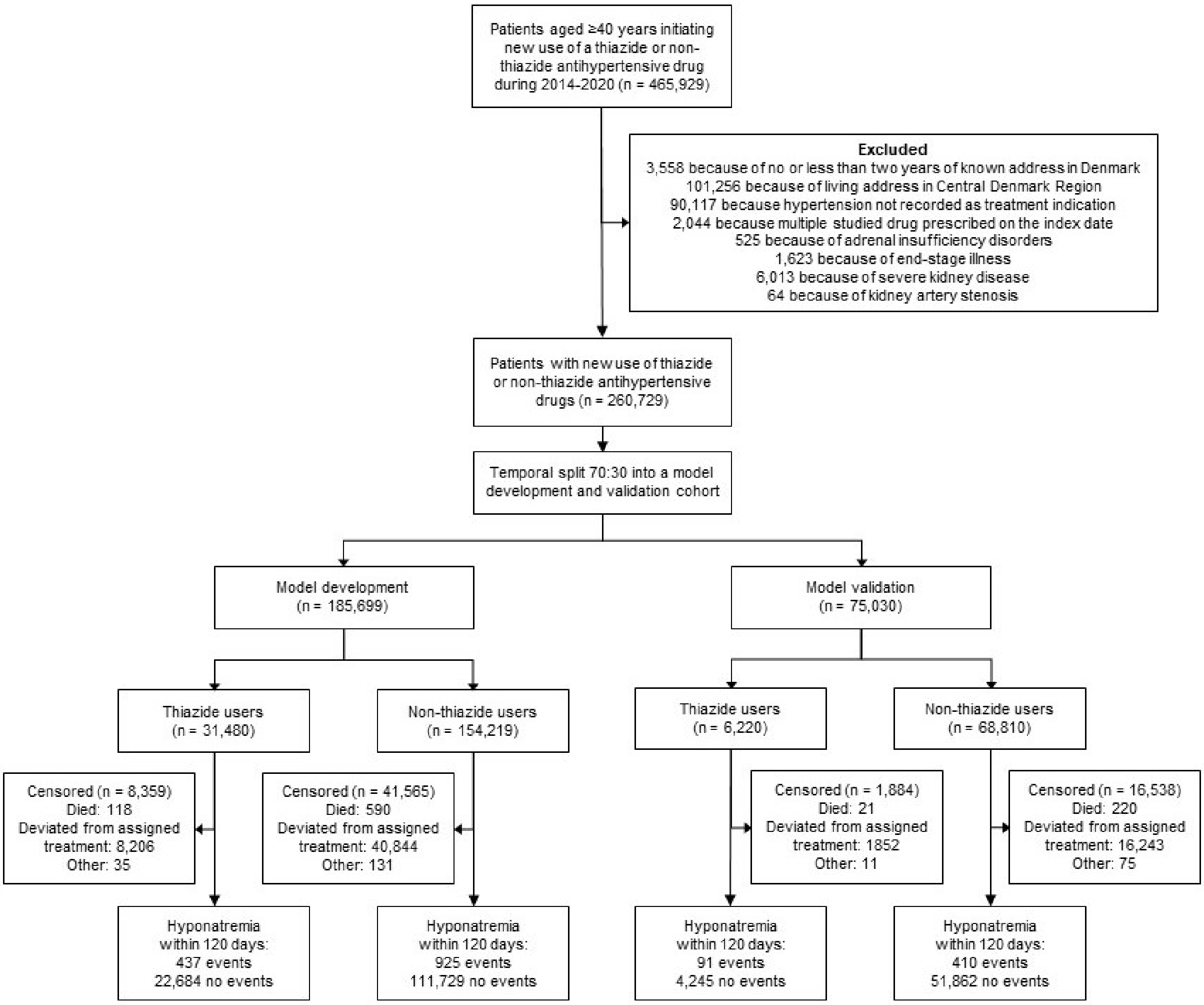

See Table S1 for definitions.

**Supplementary Table S3. Variables included and their relative variable importance in the seven selected models in the development cohort**

| Variable | Relative variable importance | | | | | | |
|---|---|---|---|---|---|---|---|
| | 1-cov | 2-cov | 3-cov | 4-cov | 5-cov | 20-cov | 66-cov (full) |
| Sodium | 1.00000 | 0.58797 | 0.50517 | 0.44733 | 0.43016 | 0.42141 | 0.23781 |
| Age | | 0.41203 | 0.31400 | 0.27025 | 0.24551 | 0.21985 | 0.16732 |
| Hemoglobin | | | 0.18082 | 0.15072 | 0.12286 | 0.06246 | 0.08905 |
| C-reactive protein | | | | 0.13170 | 0.10657 | 0.04111 | 0.05106 |
| Lactate dehydrogenase | | | | | 0.09490 | 0.02617 | 0.03427 |
| History of hyponatremia | | | | | | 0.01724 | 0.03233 |
| Thrombocytes | | | | | | 0.02470 | 0.03184 |
| Years of education | | | | | | 0.01664 | 0.02781 |
| Albumin | | | | | | 0.01320 | 0.02579 |
| Desmopressin | | | | | | 0.03361 | 0.02377 |
| Alkaline phosphatase | | | | | | 0.01590 | 0.02100 |
| Drugs used for obstipation, diarrhea, or intestinal antiinflammation/infection | | | | | | 0.01518 | 0.02088 |
| Drug abuse | | | | | | 0.01242 | 0.01981 |
| Leukocytes | | | | | | 0.01706 | 0.01871 |
| Days of hospitalization in the past year | | | | | | 0.00987 | 0.01801 |
| Malignancy associated with hyponatremia | | | | | | 0.00826 | 0.01679 |
| Carbamide | | | | | | 0.01529 | 0.01522 |
| Low-density lipoprotein-cholesterol | | | | | | 0.01345 | 0.01450 |
| Estimated glomerular filtration rate | | | | | | 0.01175 | 0.01420 |
| Dehydration | | | | | | 0.00442 | 0.01293 |
| Frail general health | | | | | | | 0.01136 |
| Antiepileptics | | | | | | | 0.01069 |
| Alanine aminotransferase | | | | | | | 0.00952 |
| Cholesterol | | | | | | | 0.00927 |
| No. of different prescription drugs in the past year | | | | | | | 0.00847 |
| Potassium | | | | | | | 0.00721 |
| Arrhythmias | | | | | | | 0.00649 |
| Household income | | | | | | | 0.00603 |
| Opioids | | | | | | | 0.00514 |
| Renal disorders | | | | | | | 0.00385 |
| Cerebrovascular disease | | | | | | | 0.00347 |
| No. of outpatient contacts in the past year | | | | | | | 0.00309 |
| Physical impairment | | | | | | | 0.00272 |
| Antacids, H2-receptor antagonists, or proton pump inhibitors | | | | | | | 0.00176 |
| Oral anticoagulants | | | | | | | 0.00166 |
| Mental impairment | | | | | | | 0.00148 |
| Alcohol abuse | | | | | | | 0.00146 |
| Sex | | | | | | | 0.00136 |
| Essential hypertension | | | | | | | 0.00134 |

**Supplementary Table S3. Variables included and their relative variable importance in the seven selected models in the development cohort**

| Variable | Relative variable importance | | | | | | |
|---|---|---|---|---|---|---|---|
| | 1-cov | 2-cov | 3-cov | 4-cov | 5-cov | 20-cov | 66-cov (full) |
| No. of primary care contacts in the past 2 years | | | | | | | 0.00131 |
| Agents for obstructive pulmonary disease | | | | | | | 0.00127 |
| Other malignancy | | | | | | | 0.00114 |
| Antipsychotics | | | | | | | 0.00113 |
| Chronic obstructive pulmonary disease | | | | | | | 0.00067 |
| Antidepressants | | | | | | | 0.00066 |
| Aspirin | | | | | | | 0.00065 |
| NSAIDs | | | | | | | 0.00061 |
| Adenosine diphosphate receptor inhibitors | | | | | | | 0.00057 |
| Liver disease and peritonitis | | | | | | | 0.00037 |
| Any malignancy | | | | | | | 0.00036 |
| Beta-blockers | | | | | | | 0.00032 |
| Lipid-lowering drugs | | | | | | | 0.00026 |
| Heart failure | | | | | | | 0.00021 |
| Rehabilitation contacts | | | | | | | 0.00021 |
| Podiatric contacts | | | | | | | 0.00018 |
| Eltroxin | | | | | | | 0.00016 |
| Other central nervous system disorders | | | | | | | 0.00011 |
| Nitrates | | | | | | | 0.00009 |
| Ischemic heart disease | | | | | | | 0.00008 |
| Diabetes | | | | | | | 0.00007 |
| Antidiabetic (not insulin) | | | | | | | 0.00005 |
| Pancreatitis | | | | | | | 0.00004 |
| Anorexia and primary polydipsia | | | | | | | 0.00000 |
| HIV | | | | | | | 0.00000 |
| Insulin | | | | | | | 0.00000 |
| Secondary hypertension | | | | | | | 0.00000 |

Variables are ordered according to variable importance in the full model (descending order). Non-dichotomous categorical variables are summed. Models are abbreviated 1-cov to 66-cov according to the number of covariates included in the respective model.

**Supplementary Table S4. Discrimination and calibration measures for the seven selected models in the development cohort**

| Models | C-for-benefit (95% CI) | Calibration regression slope-coefficient (95% CI) | Calibration regression slope-intercept (95% CI) |
|---|---|---|---|
| 1-cov | 0.603 (0.592 to 0.614) | 1.405 (1.303 to 1.507) | -0.001 (-0.003 to 0.000) |
| 2-cov | 0.651 (0.620 to 0.682) | 1.246 (1.153 to 1.340) | -0.000 (-0.002 to 0.001) |
| 3-cov | 0.647 (0.617 to 0.677) | 1.372 (1.195 to 1.548) | -0.001 (-0.004 to 0.002) |
| 4-cov | 0.639 (0.616 to 0.662) | 1.554 (1.454 to 1.654) | -0.001 (-0.003 to 0.001) |
| 5-cov | 0.633 (0.615 to 0.652) | 1.437 (1.363 to 1.510) | -0.001 (-0.002 to 0.000) |
| 20-cov | 0.623 (0.614 to 0.632) | 1.554 (1.346 to 1.762) | -0.001 (-0.004 to 0.002) |
| 66-cov (full) | 0.604 (0.592 to 0.616) | 1.298 (1.230 to 1.366) | -0.001 (-0.002 to 0.000) |

CI = confidence interval. Models are abbreviated 1-cov to 66-cov according to the number of covariates included in the respective model. The corresponding calibration plots for the slope coefficients and intercepts are shown in Figure S2.

**Supplementary Figure S2. Calibration plot for the seven selected models in the development cohort**

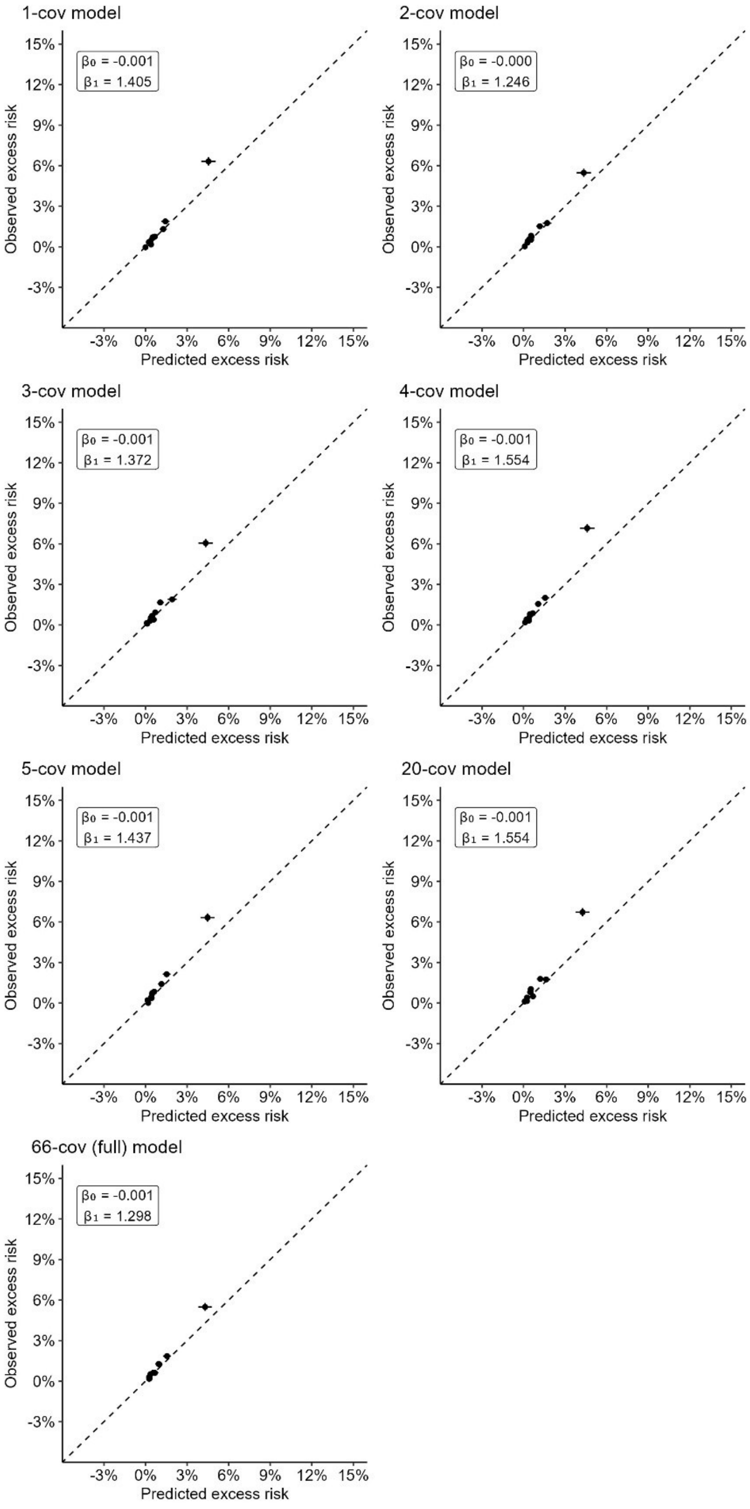

Models are abbreviated 1-cov to 66-cov according to the number of covariates included in the respective model. Thiazide and non-thiazide users within the development cohort were matched 1:1 (to obtain observed excess risks) and grouped into 10 groups according to their predicted excess risk of hyponatremia.

### Supplementary Table S5. Population benefit measures in the seven selected models by high-risk group definitions in the development cohort

| High-risk group definition and model | Average excess risk in thiazide-treated (95% CI) | Number of patients in the group[a] | Average excess risk avoided in thiazide-treated if high-risk group not treated with thiazide (95% CI) | Number needed to not treat with thiazide in the group to prevent 1 TIH case (95% CI) |
|---|---|---|---|---|
| **All** | | | | |
| 1-cov | 1.0% (0.6% to 1.4%) | 31604 | NA | 104 (175 to 74) |
| 2-cov | 1.0% (0.6% to 1.4%) | 31604 | NA | 104 (175 to 74) |
| 3-cov | 1.0% (0.6% to 1.4%) | 31604 | NA | 104 (178 to 73) |
| 4-cov | 1.0% (0.6% to 1.4%) | 31604 | NA | 104 (177 to 73) |
| 5-cov | 1.0% (0.6% to 1.4%) | 31604 | NA | 104 (178 to 73) |
| 20-cov | 1.0% (0.6% to 1.4%) | 31604 | NA | 104 (178 to 73) |
| 66-cov (full) | 1.0% (0.6% to 1.4%) | 31604 | NA | 104 (176 to 73) |
| **10% of thiazide-treated with the highest excess risk** | | | | |
| 1-cov | 3.6% (1.4% to 5.7%) | 3421 | 0.3% (-0.1% to 0.8%) | 28 (68 to 18) |
| 2-cov | 3.6% (0.0% to 6.7%) | 3163 | 0.3% (-0.2% to 0.8%) | 28 (158 to 15) |
| 3-cov | 3.9% (0.1% to 7.1%) | 3163 | 0.3% (-0.2% to 0.8%) | 26 (121 to 15) |
| 4-cov | 4.1% (1.2% to 7.0%) | 3162 | 0.3% (-0.1% to 0.8%) | 25 (78 to 15) |
| 5-cov | 4.0% (1.2% to 6.8%) | 3163 | 0.3% (-0.1% to 0.8%) | 25 (79 to 15) |
| 20-cov | 3.9% (1.1% to 6.7%) | 3162 | 0.3% (-0.1% to 0.8%) | 26 (81 to 15) |
| 66-cov (full) | 4.4% (1.9% to 6.8%) | 3162 | 0.4% (-0.1% to 0.8%) | 23 (52 to 15) |
| **5% of thiazide-treated with the highest excess risk** | | | | |
| 1-cov | 5.8% (1.3% to 10.4%) | 1717 | 0.2% (-0.3% to 0.7%) | 22 (128 to 12) |
| 2-cov | 4.6% (-1.4% to 10.6%) | 1583 | 0.2% (-0.3% to 0.7%) | 23 (-281 to 11) |
| 3-cov | 4.8% (-1.2% to 10.8%) | 1582 | 0.2% (-0.3% to 0.7%) | 22 (-247 to 11) |
| 4-cov | 5.5% (0.4% to 10.6%) | 1582 | 0.2% (-0.3% to 0.7%) | 20 (205 to 11) |
| 5-cov | 5.6% (0.5% to 10.7%) | 1582 | 0.2% (-0.3% to 0.7%) | 21 (240 to 11) |
| 20-cov | 4.9% (-0.1% to 10.0%) | 1582 | 0.2% (-0.3% to 0.7%) | 22 (238 to 11) |
| 66-cov (full) | 6.1% (2.0% to 10.2%) | 1582 | 0.2% (-0.3% to 0.7%) | 19 (77 to 11) |

CI = confidence interval, NA = not applicable, TIH = thiazide-induced hyponatremia. Models are abbreviated 1-cov to 66-cov according to the number of covariates included in the respective model.
[a]The group consists of treated patients from the full development cohort, including censored patients. The reported size is the total IPCW weight from non-censored treated patients in the group.

**Supplementary Figure S3. Calibration plot for the seven selected models in the validation cohort**

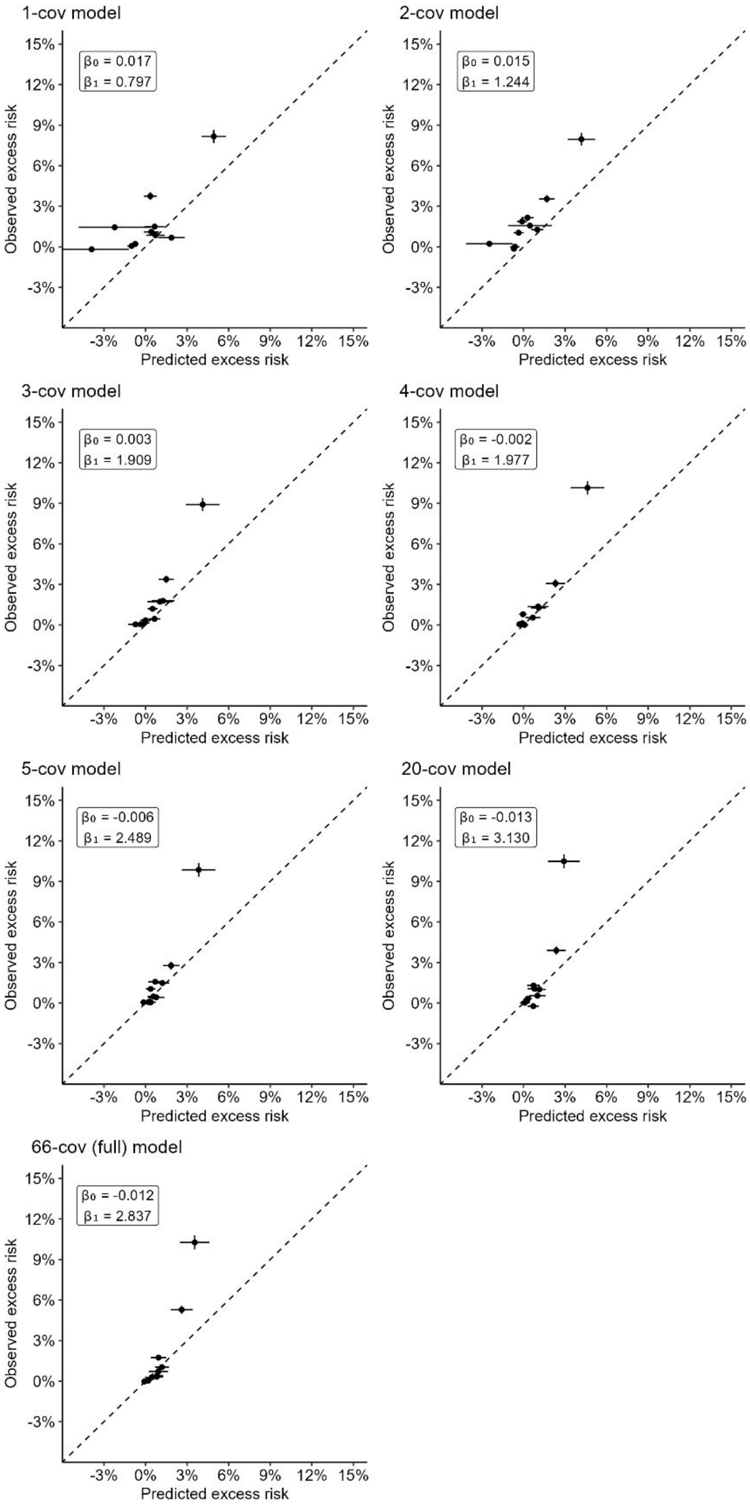

Models are abbreviated 1-cov to 66-cov according to the number of covariates included in the respective model. Thiazide and non-thiazide users within the validation cohort were matched 1:1 (to obtain observed excess risks) and grouped into 10 groups according to their predicted excess risk of hyponatremia.

**Supplementary Table S6. Sensitivity analysis of calibration for the seven selected models in the validation cohort grouping patients using five and twenty groups (instead of ten groups used in the main analyses) according to predicted excess risk of hyponatremia**

| Alternative grouping and models | Calibration regression slope-coefficient (95% CI) | Calibration regression slope-intercept (95% CI) |
|---|---|---|
| **Five groups** | | |
| 1-cov | 0.871 (0.186 to 1.557) | 0.018 (0.004 to 0.033) |
| 2-cov | 1.190 (0.667 to 1.713) | 0.016 (0.007 to 0.024) |
| 3-cov | 1.681 (0.512 to 2.850) | 0.005 (-0.012 to 0.022) |
| 4-cov | 2.081 (1.184 to 2.979) | -0.002 (-0.015 to 0.012) |
| 5-cov | 2.491 (2.020 to 2.961) | -0.008 (-0.015 to -0.002) |
| 20-cov | 3.152 (0.892 to 5.413) | -0.014 (-0.043 to 0.016) |
| 66-cov (full) | 2.916 (1.111 to 4.721) | -0.013 (-0.040 to 0.014) |
| **Twenty groups** | | |
| 1-cov | 0.388 (0.127 to 0.649) | 0.019 (0.007 to 0.030) |
| 2-cov | 0.757 (0.396 to 1.118) | 0.016 (0.007 to 0.025) |
| 3-cov | 1.601 (1.093 to 2.109) | 0.005 (-0.003 to 0.013) |
| 4-cov | 2.005 (1.237 to 2.773) | -0.001 (-0.013 to 0.011) |
| 5-cov | 2.149 (1.449 to 2.850) | -0.005 (-0.017 to 0.006) |
| 20-cov | 2.141 (0.815 to 3.467) | -0.003 (-0.022 to 0.015) |
| 66-cov (full) | 2.779 (1.755 to 3.803) | -0.009 (-0.024 to 0.005) |

CI = confidence intervals. Models are abbreviated 1-cov to 66-cov according to the number of covariates included in the respective model. The corresponding calibration plots of the slope coefficients and intercepts are shown in Figure S4 (for five groups) and S5 (for twenty groups).

**Supplementary Figure S4. Sensitivity analysis of calibration plots for the seven selected models in the validation cohort grouping in five groups according to predicted excess risk**

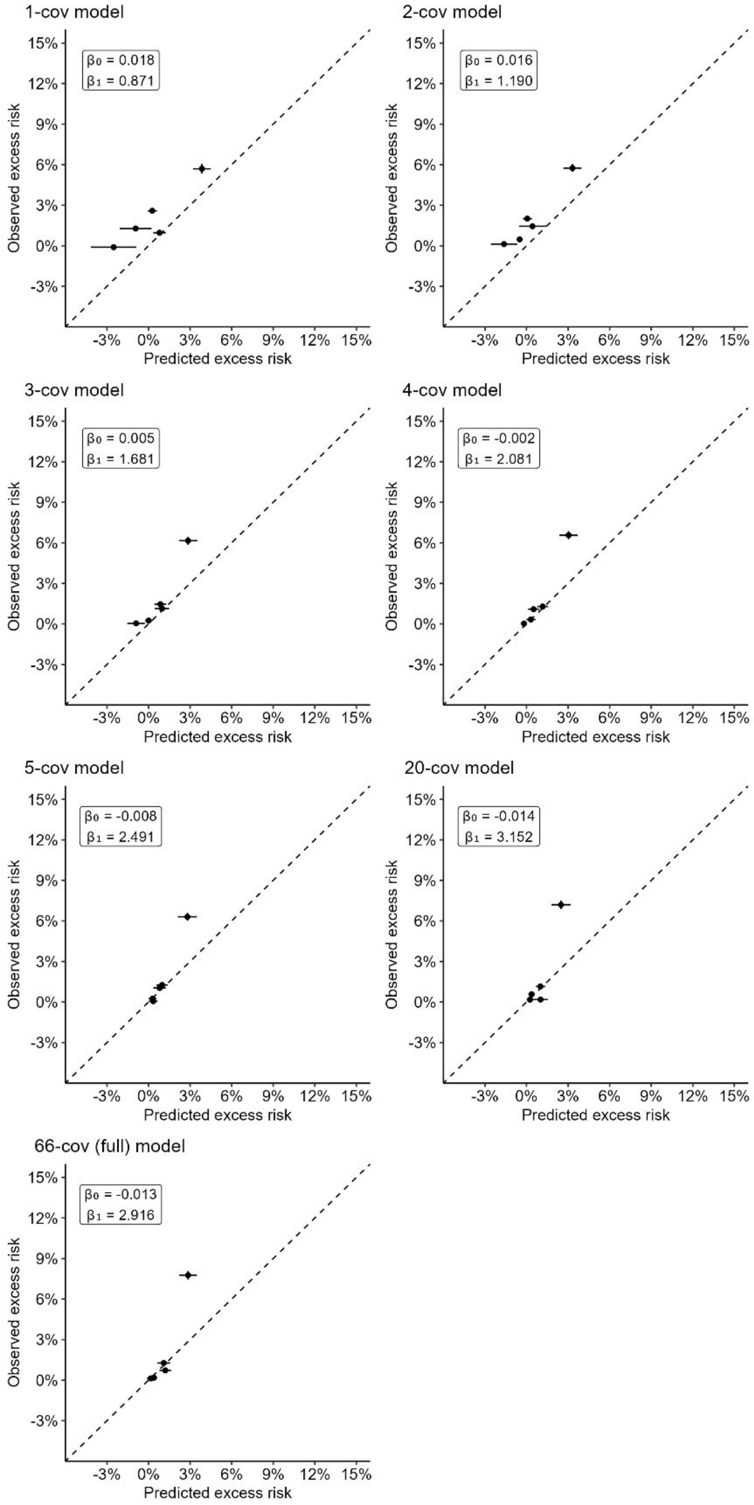

Models are abbreviated 1-cov to 66-cov according to the number of covariates included in the respective model. In this sensitivity analysis, thiazide and non-thiazide users within the validation cohort were matched 1:1 (to obtain observed excess risks) and grouped into five groups according to their predicted excess risk of hyponatremia.

**Supplementary Figure S5. Sensitivity analysis of calibration plots for the seven selected models in the validation cohort grouping in twenty groups according to predicted excess risk**

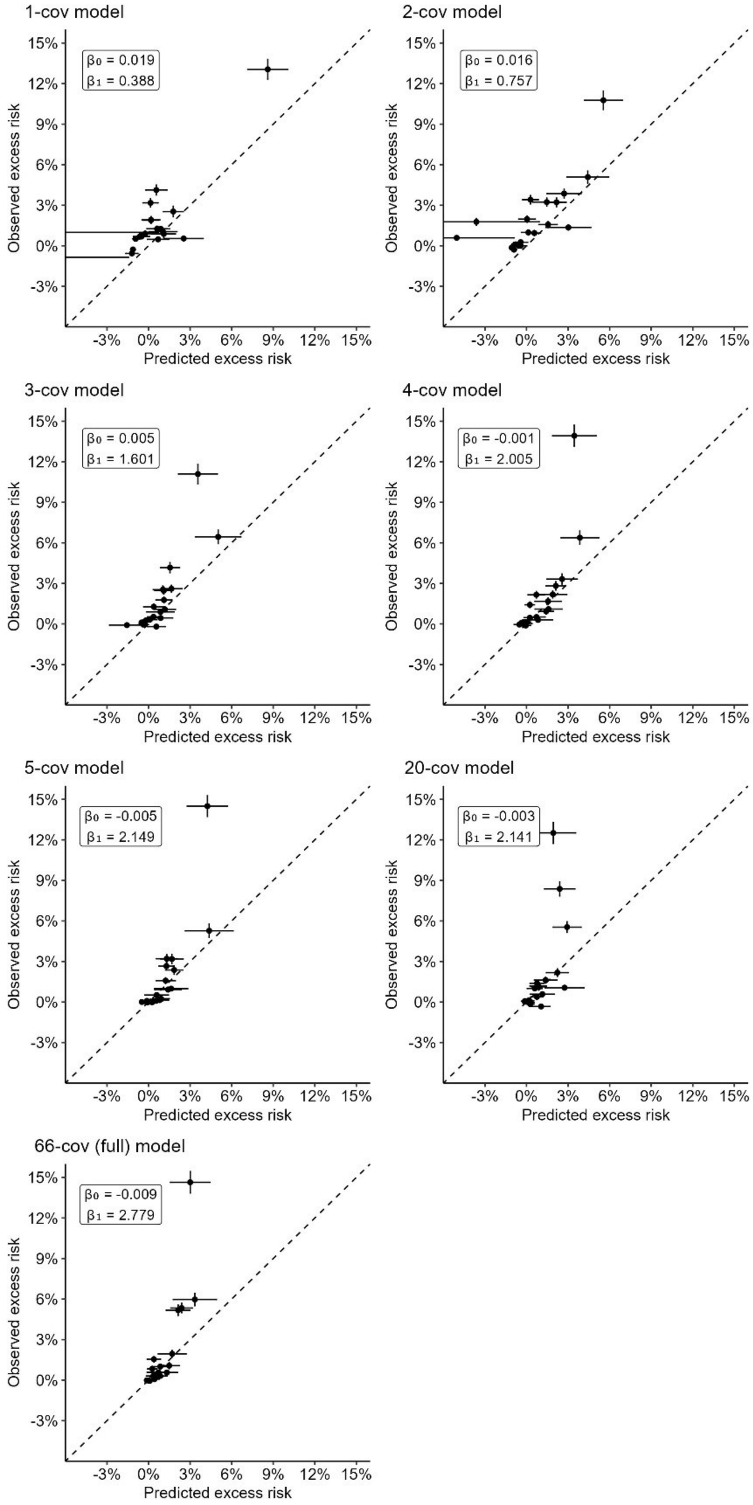

Models are abbreviated 1-cov to 66-cov according to the number of covariates included in the respective model. In this sensitivity analysis, thiazide and non-thiazide users within the validation cohort were matched 1:1 (to obtain observed excess risks) and grouped into twenty groups according to their predicted excess risk of hyponatremia.

**Supplementary Figure S6. Agreement on the identification of high-risk (and not high risk) patients between the seven selected models by high-risk group definitions in the validation cohort**

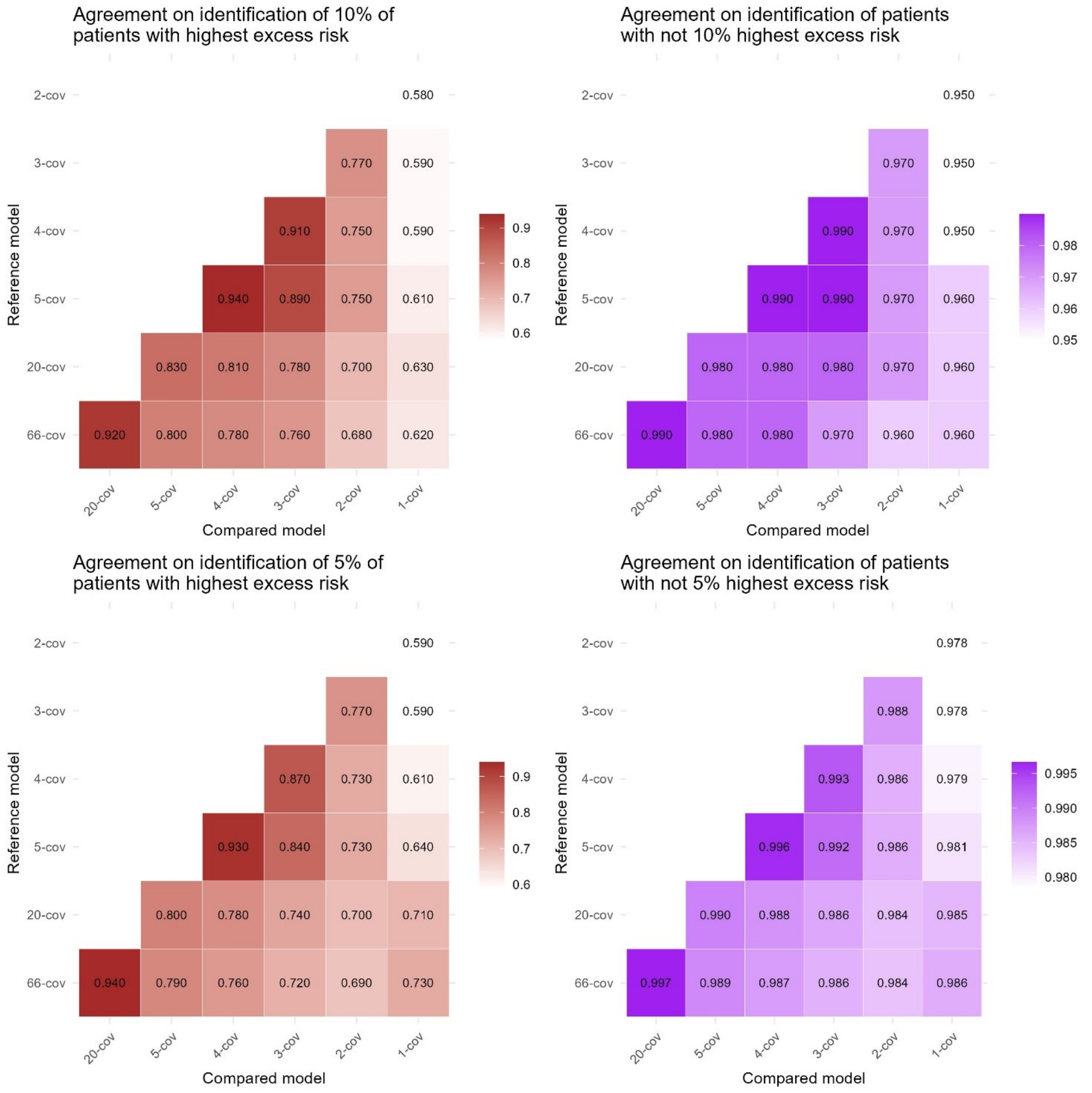

Models are abbreviated 1-cov to 66-cov according to the number of covariates included in the respective model. Agreement between the selected models with respect to identifying these high-risk (and non-high risk) patients was assessed by alternately treating each model as the gold standard to calculate the proportion of agreement between the high-risk group of each other model and the high-risk group of the gold standard model.

### Supplementary Figure S7. Correlation of individual excess risk between the seven selected models in the validation cohort

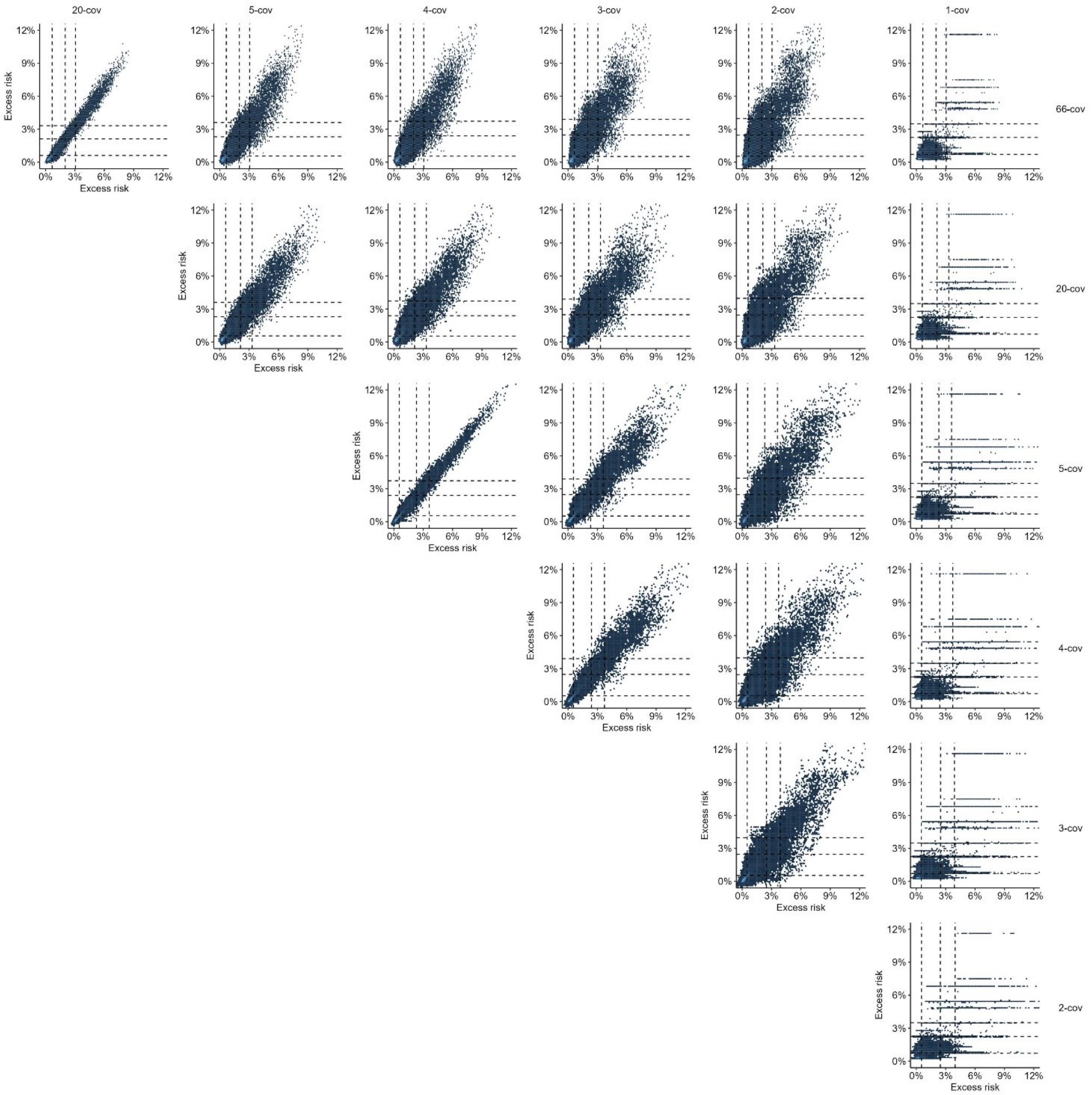

Models are abbreviated 1-cov to 66-cov according to the number of covariates included in the respective model. Columns are compared model (y-axis), and rows are reference model (x-axis). Correlation coefficients are provided in Table S7. Vertical (from bottom to top) and horizontal (from left to right) dashed lines represent cohort centile proportions of 50%, 90%, and 95% according to the respective compared models.

**Supplementary Table S7. Correlation coefficients of predicted excess risk between the seven selected models in the validation cohort**

| **20-cov** | **5-cov** | **4-cov** | **3-cov** | **2-cov** | **1-cov** | |
|---|---|---|---|---|---|---|
| 0.98 | 0.93 | 0.91 | 0.88 | 0.84 | 0.75 | **66-cov** |
| | 0.94 | 0.93 | 0.90 | 0.86 | 0.75 | **20-cov** |
| | | 0.99 | 0.96 | 0.90 | 0.72 | **5-cov** |
| | | | 0.97 | 0.90 | 0.70 | **4-cov** |
| | | | | 0.91 | 0.69 | **3-cov** |
| | | | | | 0.69 | **2-cov** |

Correlation coefficients correspond to the comparisons of Figure S7. Correlation coefficients are Pearson's correlation coefficients.

### Supplementary Figure S8. Average excess risk and distribution of individual excess risk by baseline variables in the validation cohort

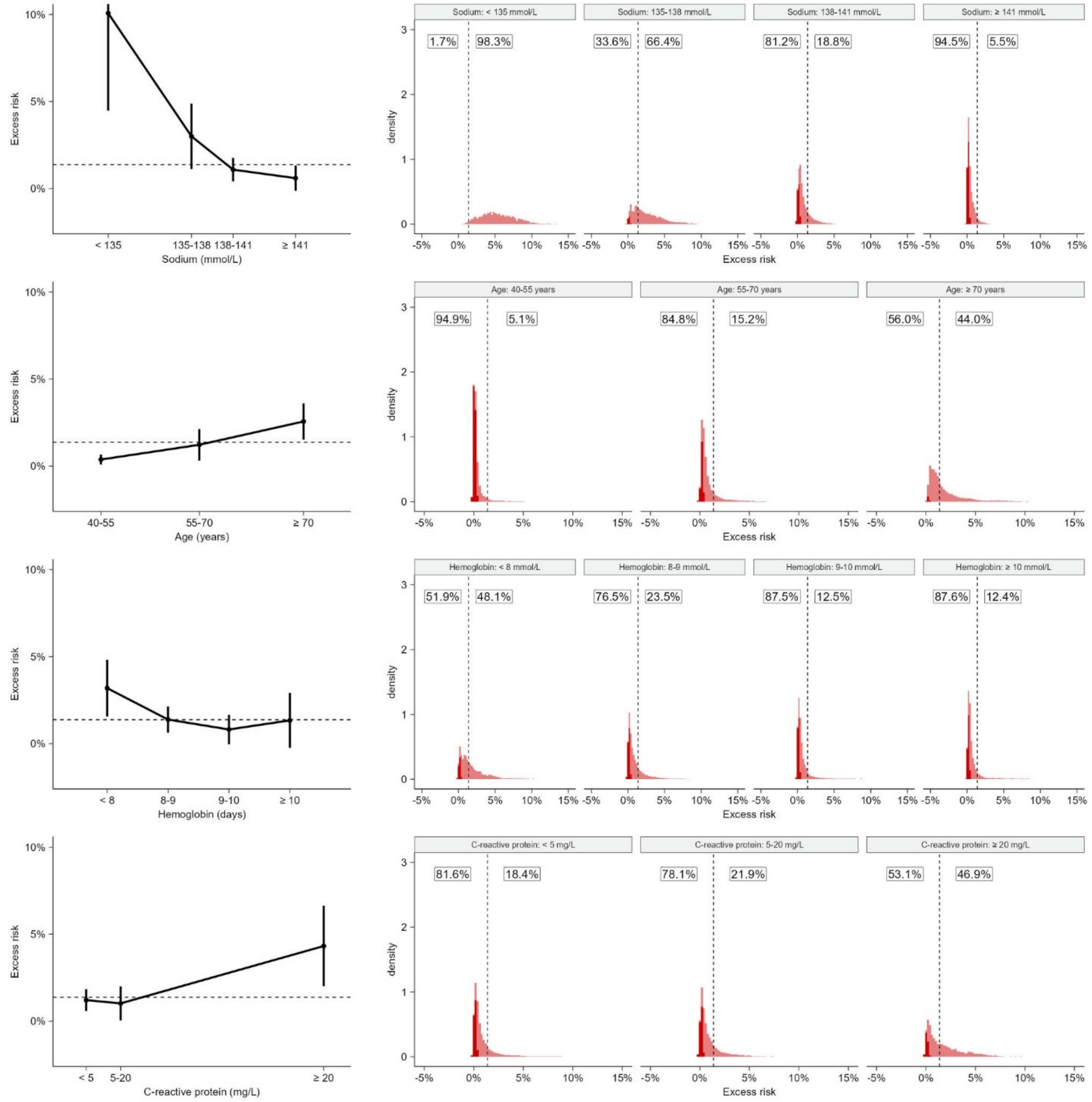

The individual-level excess risk predictions are based on the four-covariate model within the validation cohort. The left column presents the excess risk of hyponatremia with thiazide use for subgroups defined by intervals of the four variables included in the four-covariate model (baseline plasma sodium, hemoglobin, and C-reactive protein levels and age). The dashed line is the average excess risk in the full population. The right column presents the distributions of the individual-level excess risk within these covariate-defined subgroups. The dashed line is the average excess risk in the full population. Depicted percentage numbers are the proportion of individuals with excess risk below or above the population average. Dark red indicates that the estimated excess risks significantly differ from the average excess risk in the full population at a 5% significance level using a z-test with a two-sided alternative.

**Supplementary Figure S9. Distributions of excess risk by baseline sodium level and age in the validation cohort**

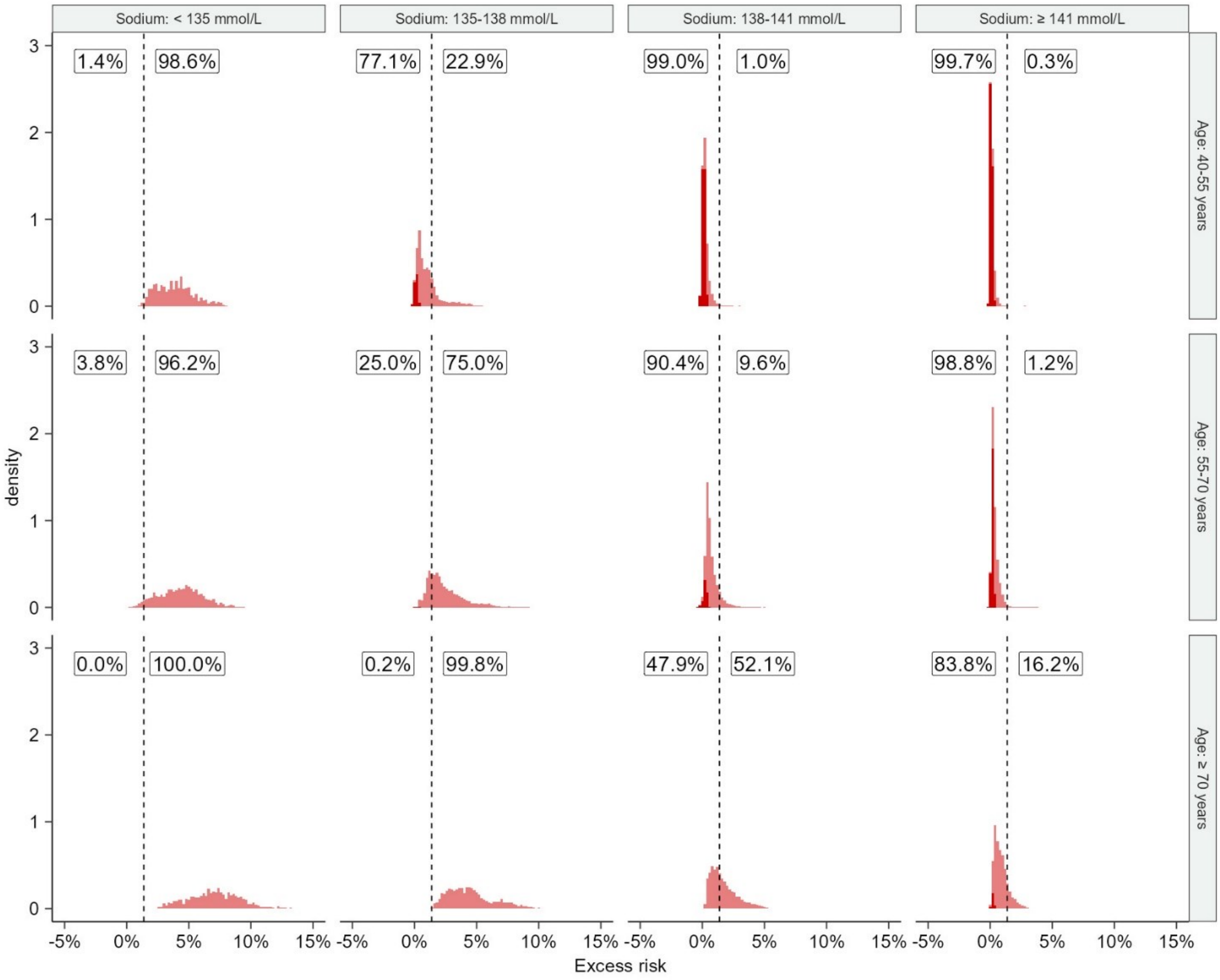

The individual-level excess risk predictions are based on the four-covariate model within the validation cohort. The figure presents the distributions of the individual-level excess risk within combinations of baseline sodium levels (columns) and age-defined (rows) subgroups. The dashed line is the average excess risk in the full population. Depicted percentage numbers are the proportion of individuals with excess risk below or above the population average. Dark red indicates that the estimated excess risks significantly differ from the population average at a 5% significance level using a z-test with a two-sided alternative.

**Supplementary Figure S10. Distributions of excess risk by baseline hemoglobin and C-reactive protein levels in the validation cohort**

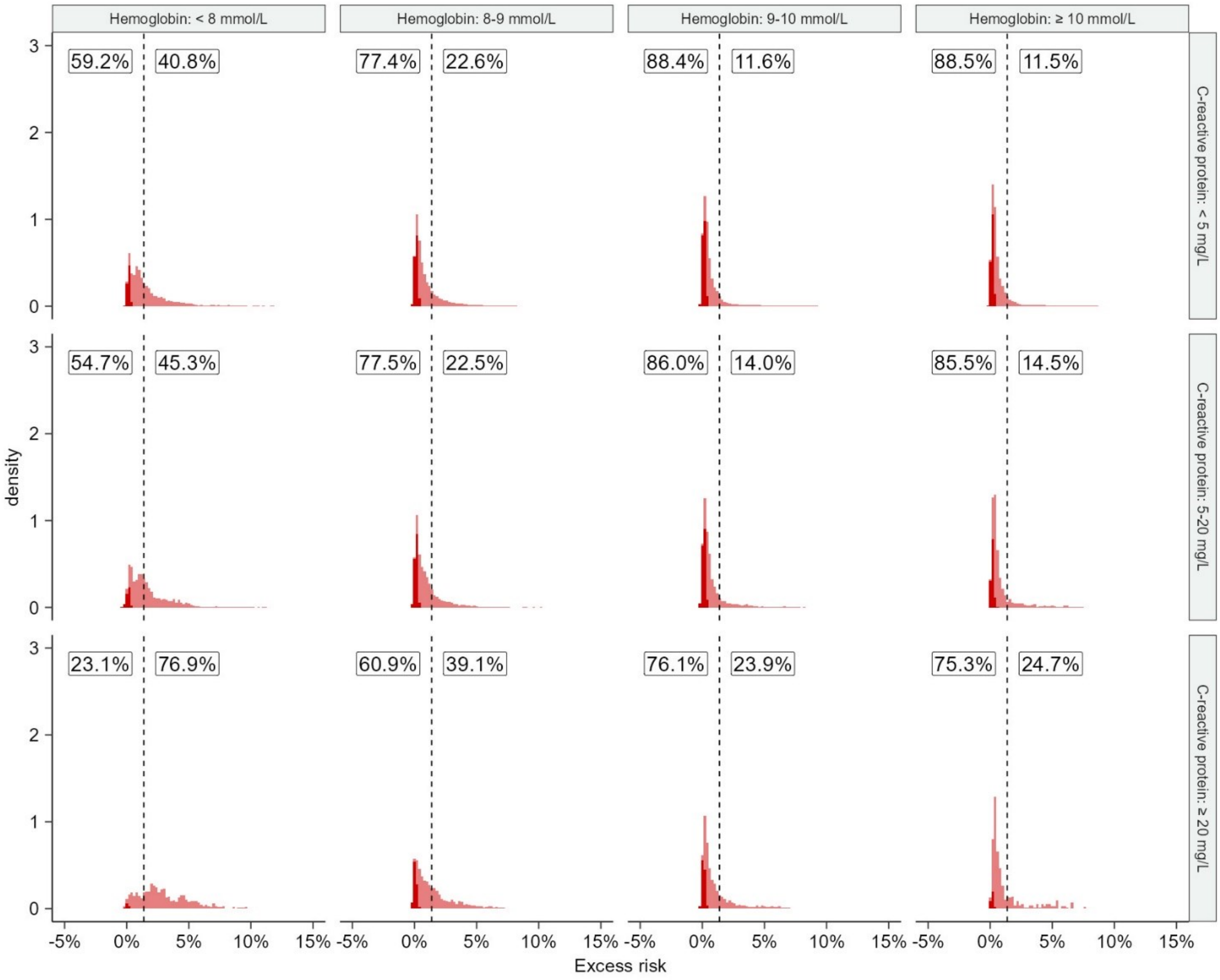

The individual-level excess risk predictions are based on the four-covariate model within the validation cohort. The figure presents the distributions of the individual-level excess risk within combinations of baseline C-reactive protein (columns) and hemoglobin level-defined (rows) subgroups. The dashed line is the average excess risk in the full population. Depicted percentage numbers are the proportion of individuals with excess risk below or above the population average. Dark red indicates that the estimated excess risks significantly differ from the population average at a 5% significance level using a z-test with a two-sided alternative.

**Supplementary Table S8. Population benefit measures in the seven selected models by high-risk group definitions in the validation cohort**

| High-risk group definition and model | Average excess risk in thiazide-treated (95% CI) | Number of patients in the group[a] | Average excess risk avoided in thiazide-treated if high-risk group not treated with thiazide (95% CI) | Number needed to not treat with thiazide in the group to prevent 1 TIH case (95% CI) |
|---|---|---|---|---|
| **All** | | | | |
| 1-cov | 1.8% (1.4% to 2.2%) | 6611 | NA | 56 (74 to 45) |
| 2-cov | 1.8% (1.4% to 2.2%) | 6611 | NA | 56 (74 to 45) |
| 3-cov | 1.8% (1.3% to 2.2%) | 6611 | NA | 56 (74 to 45) |
| 4-cov | 1.8% (1.3% to 2.2%) | 6611 | NA | 56 (74 to 45) |
| 5-cov | 1.8% (1.3% to 2.2%) | 6611 | NA | 56 (74 to 45) |
| 20-cov | 1.8% (1.3% to 2.2%) | 6611 | NA | 56 (74 to 45) |
| 66-cov (full) | 1.8% (1.3% to 2.2%) | 6611 | NA | 56 (74 to 45) |
| **10% of thiazide-treated with the highest excess risk** | | | | |
| 1-cov | 5.3% (3.1% to 7.4%) | 914 | 0.6% (0.0% to 1.2%) | 19 (32 to 13) |
| 2-cov | 7.6% (4.6% to 10.5%) | 664 | 0.7% (0.1% to 1.2%) | 13 (22 to 10) |
| 3-cov | 7.0% (4.0% to 10.0%) | 664 | 0.6% (0.1% to 1.2%) | 14 (25 to 10) |
| 4-cov | 7.4% (4.4% to 10.5%) | 664 | 0.7% (0.1% to 1.2%) | 13 (23 to 10) |
| 5-cov | 8.0% (4.8% to 11.1%) | 663 | 0.8% (0.2% to 1.3%) | 13 (21 to 9) |
| 20-cov | 8.0% (4.8% to 11.2%) | 664 | 0.8% (0.3% to 1.3%) | 12 (21 to 9) |
| 66-cov (full) | 8.6% (5.4% to 11.9%) | 664 | 0.9% (0.4% to 1.4%) | 12 (19 to 8) |
| **5% of thiazide-treated with the highest excess risk** | | | | |
| 1-cov | 8.7% (4.8% to 12.6%) | 473 | 0.6% (0.0% to 1.1%) | 11 (21 to 8) |
| 2-cov | 9.9% (5.0% to 14.8%) | 333 | 0.5% (-0.1% to 1.0%) | 10 (20 to 7) |
| 3-cov | 7.7% (3.0% to 12.4%) | 333 | 0.3% (-0.2% to 0.9%) | 13 (33 to 8) |
| 4-cov | 9.6% (4.8% to 14.5%) | 333 | 0.5% (-0.0% to 1.1%) | 10 (21 to 7) |
| 5-cov | 10.0% (4.9% to 15.0%) | 333 | 0.5% (-0.0% to 1.1%) | 10 (20 to 7) |
| 20-cov | 11.3% (6.0% to 16.6%) | 334 | 0.6% (0.0% to 1.1%) | 9 (16 to 6) |
| 66-cov (full) | 11.0% (5.6% to 16.3%) | 333 | 0.6% (0.0% to 1.1%) | 9 (18 to 6) |

CI = confidence interval, TIH = thiazide-induced hyponatremia. Models are abbreviated 1-cov to 66-cov according to the number of covariates included in the respective model.

[a]The group consists of treated patients from the full validation cohort, including censored patients. The reported size is the total IPCW weight from non-censored treated patients in the group.

**Supplementary Table S9. Discrimination and calibration measures in the development and the validation cohort for each of the two thiazide subtypes separately**

| Cohort and thiazide subtype | C-for-benefit (95% CI) | Calibration regression slope-coefficient (95% CI) | Calibration regression slope-intercept (95% CI) |
|---|---|---|---|
| **Development cohort** | | | |
| Bendroflumethiazide | 0.644 (0.623 to 0.665) | 1.696 (1.520 to 1.871) | -0.004 (-0.006 to -0.001) |
| Hydrochlorothiazide | 0.661 (0.650 to 0.673) | 1.504 (1.353 to 1.656) | -0.003 (-0.006 to 0.000) |
| **Validation cohort** | | | |
| Bendroflumethiazide | 0.631 (0.622 to 0.641) | 2.089 (1.424 to 2.753) | -0.007 (-0.020 to 0.005) |
| Hydrochlorothiazide | 0.716 (0.710 to 0.722) | 0.340 (-0.210 to 0.891) | 0.030 (-0.006 to 0.066) |

All measures were estimated using four-covariate models. In the development cohort, the model for bendroflumethiazide use was derived based on 17,904 users of the thiazide vs 51,240 calcium-channel blocker users, and the hydrochlorothiazide model was based on 13,576 users of the thiazide vs 102,979 renin-angiotensin-system inhibitor users (the hydrochlorothiazide-containing pills are combinatory pills also containing a renin-angiotensin-system inhibitor; see Table S1 for definitions). Corresponding numbers in the validation cohort were 4,088 vs 22,316 and 2,132 vs 46,494 users, respectively.

**Supplementary Figure S11. Calibration plot in the development and the validation cohort for each of the two thiazide subtypes separately**

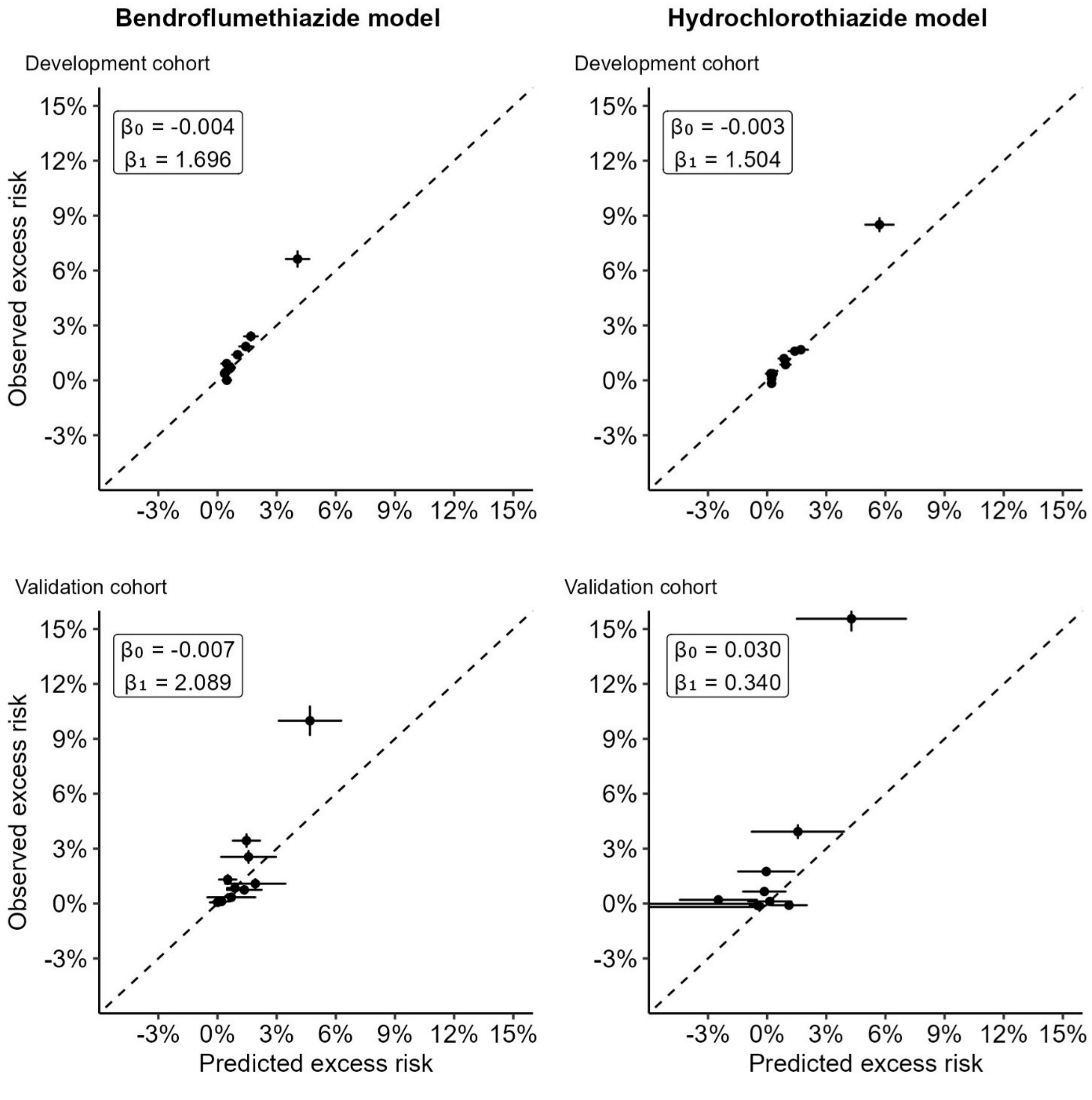

All measures were estimated using four-covariate models. Thiazide and non-thiazide users were matched 1:1 (to obtain observed excess risks) and grouped into 10 groups according to their predicted excess risk of hyponatremia.

**Supplementary Table S10. Population benefit measures in the development and the validation cohort for each of the two thiazide subtypes separately by high-risk group definitions**

| Cohort, high-risk group definition and thiazide subtype | Average excess risk in thiazide-treated (95% CI) | Average excess risk avoided in thiazide-treated if high-risk group not treated with thiazide (95% CI) | Number needed to not treat with thiazide in the group to prevent 1 TIH case (95% CI) |
|---|---|---|---|
| **Development cohort** | | | |
| **All** | | | |
| Bendroflumethiazide | 1.0% (0.6% to 1.4%) | NA | 100 (164 to 72) |
| Hydrochlorothiazide | 1.0% (0.6% to 1.5%) | NA | 96 (171 to 67) |
| **10% of thiazide-treated with the highest excess risk** | | | |
| Bendroflumethiazide | 4.0% (1.3% to 6.8%) | 0.3% (-0.2% to 0.8%) | 25 (63 to 15) |
| Hydrochlorothiazide | 4.5% (1.8% to 7.2%) | 0.4% (-0.2% to 0.9%) | 22 (51 to 14) |
| **5% of thiazide-treated with the highest excess risk** | | | |
| Bendroflumethiazide | 4.9% (0.3% to 9.4%) | 0.2% (-0.3% to 0.7%) | 21 (126 to 11) |
| Hydrochlorothiazide | 5.9% (1.2% to 10.5%) | 0.3% (-0.3% to 0.8%) | 17 (53 to 10) |
| **Validation cohort** | | | |
| **All** | | | |
| Bendroflumethiazide | 1.6% (1.0% to 2.1%) | NA | 63 (96 to 47) |
| Hydrochlorothiazide | 2.3% (1.5% to 3.1%) | NA | 44 (67 to 33) |
| **10% of thiazide-treated with the highest excess risk** | | | |
| Bendroflumethiazide | 6.6% (2.9% to 10.2%) | 0.6% (-0.1% to 1.3%) | 15 (34 to 10) |
| Hydrochlorothiazide | 11.2% (5.6% to 16.7%) | 1.1% (0.1% to 2.0%) | 9 (18 to 6) |
| **5% of thiazide-treated with the highest excess risk** | | | |
| Bendroflumethiazide | 8.3% (2.3% to 14.2%) | 0.4% (-0.3% to 1.1%) | 12 (42 to 7) |
| Hydrochlorothiazide | 14.1% (4.9% to 23.2%) | 0.7% (-0.3% to 1.7%) | 7 (20 to 4) |

NA = not applicable. All measures were estimated using four-covariate models. The bendroflumethiazide model was derived based on 17,904 users of the thiazide vs 51,240 calcium-channel blocker users, and the hydrochlorothiazide model was based on 13,576 users of the thiazide vs 102,979 renin-angiotensin-system inhibitor users (the hydrochlorothiazide-containing pills are combinatory pills also containing a renin-angiotensin-system inhibitor; see Table S1 for definitions). Corresponding numbers in the validation cohort were 4,088 vs 22,316 and 2,132 vs 46,494 users, respectively.

**Supplementary Table S11. Sensitivity analysis for discrimination and calibration measures handling missing covariates in the validation cohort.**

| Handling of missingness | C-for-benefit (95% CI) | Calibration regression slope-coefficient (95% CI) | Calibration regression slope-intercept (95% CI) |
|---|---|---|---|
| Complete-case | 0.649 (0.638 to 0.659) | 2.520 (1.074 to 3.967) | -0.007 (-0.025 to 0.012) |
| Multiple-imputation | 0.659 (0.652 to 0.666) | 2.069 (0.150 to 3.989) | -0.002 (-0.024 to 0.020) |

All measures were estimated using four-covariate models. The model with complete-case handling of missingness was derived based on 18,467 thiazide vs 100,010 non-thiazide users. Measures for the model with complete-case handling of missingness were estimated in the validation cohort with 5,046 thiazide vs 61,094 non-thiazide users, respectively (measures for the multiple imputation handling model were estimated in the main validation cohort). Included laboratory blood test results were restricted to those with missingness <10% in the validation cohort for both the complete-case and the compared multiple-imputation model to preserve sample sizes.

**Supplementary Table S12. Sensitivity analysis for population benefit measures handling missing covariates in the validation cohort**

| Model and high-risk group definition | Average excess risk in thiazide-treated (95% CI) | Average excess risk avoided in thiazide-treated if high-risk group not treated with thiazide (95% CI) | Number needed to not treat with thiazide in the group to prevent 1 TIH case (95% CI) |
|---|---|---|---|
| **All** | | | |
| Complete-case model | 1.7% (1.2% to 2.1%) | NA | 60 (83 to 48) |
| Multiple-imputation model | 1.6% (1.2% to 2.0%) | NA | 61 (82 to 49) |
| **10% of thiazide-treated with the highest excess risk** | | | |
| Complete-case model | 8.8% (5.6% to 12.1%) | 0.8% (0.3% to 1.4%) | 11 (18 to 8) |
| Multiple-imputation model | 8.8% (5.6% to 12.0%) | 0.9% (0.4% to 1.4%) | 11 (18 to 8) |
| **5% of thiazide-treated with the highest excess risk** | | | |
| Complete-case model | 11.6% (6.2% to 17.0%) | 0.6% (-0.0% to 1.1%) | 9 (16 to 6) |
| Multiple-imputation model | 10.2% (4.9% to 15.5%) | 0.5% (-0.0% to 1.0%) | 10 (20 to 6) |

NA = not applicable. The model with complete-case handling of missingness was derived based on 18,467 thiazide vs 100,010 non-thiazide users. Measures for the model with complete-case handling of missingness were estimated in the validation cohort with 5,046 thiazide vs 61,094 non-thiazide users, respectively (measures for the multiple imputation handling model were estimated in the main validation cohort). Included laboratory blood test results were restricted to those with missingness <10% in the validation cohort for both the complete-case and the compared multiple-imputation model to preserve sample sizes.

**Supplementary Table S13. Average excess risk of hyponatremia with thiazide use in stratified subgroups of a priori chosen variables in the development and validation cohorts combined**

| Variable | Subgroup | N | Average excess risk (95% CI) |
|---|---|---|---|
| Household income | Q1 | 25816 | 0.74 (0.32 to 1.15) |
| | Q2 | 42213 | 1.04 (0.68 to 1.40) |
| | Q3 | 43050 | 1.09 (0.67 to 1.51) |
| | Q4 | 42632 | 1.48 (1.04 to 1.92) |
| | Q5 | 38440 | 0.84 (0.42 to 1.27) |
| Heart failure | Yes | 843 | 0.30 (-2.14 to 2.75) |
| | No | 191310 | 1.07 (0.90 to 1.25) |
| Ischemic heart disease | Yes | 8280 | 1.46 (0.68 to 2.24) |
| | No | 183873 | 1.05 (0.87 to 1.23) |
| Cerebrovascular disease | Yes | 9899 | 2.19 (1.04 to 3.34) |
| | No | 182254 | 1.01 (0.83 to 1.18) |
| Any malignancy | Yes | 10156 | 1.52 (0.66 to 2.39) |
| | No | 181997 | 1.04 (0.86 to 1.22) |
| Malignancy association with hyponatremia | Yes | 6675 | 2.27 (1.05 to 3.48) |
| | No | 185478 | 1.02 (0.85 to 1.20) |
| Other malignancy | Yes | 8925 | 1.77 (0.80 to 2.73) |
| | No | 183228 | 1.03 (0.86 to 1.21) |
| Liver disease, peritonitis, or pancreatitis | Yes | 2233 | 0.74 (-1.07 to 2.55) |
| | No | 189920 | 1.07 (0.90 to 1.25) |
| COPD and COPD drug use | Yes | 21693 | 1.53 (0.96 to 2.09) |
| | No | 170460 | 1.01 (0.82 to 1.20) |
| Diabetes and antidiabetic drug use | Yes | 10755 | 1.06 (0.35 to 1.78) |
| | No | 173711 | 1.05 (0.86 to 1.24) |
| Dehydration | Yes | 1281 | 4.58 (1.63 to 7.52) |
| | No | 190872 | 1.04 (0.87 to 1.22) |
| Frailty conditions | Yes | 25843 | 1.70 (1.20 to 2.20) |
| | No | 161309 | 0.91 (0.72 to 1.10) |
| Alcohol abuse | Yes | 3356 | 1.38 (-0.48 to 3.24) |
| | No | 188797 | 1.06 (0.89 to 1.24) |
| Prior hyponatremia | never | 188978 | 0.96 (0.79 to 1.13) |
| | ≥4 months | 1228 | 6.63 (2.69 to 10.56) |
| | <4 months | 1947 | 8.15 (2.12 to 14.18) |
| Opioids | Yes | 21999 | 1.84 (1.25 to 2.43) |
| | No | 170154 | 0.97 (0.78 to 1.15) |
| Antidepressants | Yes | 21186 | 1.37 (0.86 to 1.87) |
| | No | 170967 | 1.03 (0.84 to 1.22) |
| Antipsychotics | Yes | 5827 | 2.24 (1.03 to 3.44) |
| | No | 186326 | 1.03 (0.85 to 1.21) |
| Sodium level | < 135 mmol/L | 4991 | 6.91 (4.09 to 9.72) |
| | 135-138 mmol/L | 19425 | 3.03 (2.17 to 3.89) |
| | 138-141 mmol/L | 79140 | 0.93 (0.66 to 1.19) |

**Supplementary Table S13. Average excess risk of hyponatremia with thiazide use in stratified subgroups of a priori chosen variables in the development and validation cohorts combined**

| Variable | Subgroup | N | Average excess risk (95% CI) |
|---|---|---|---|
| | ≥ 141 mmol/L | 88594 | 0.42 (0.21 to 0.63) |
| eGFR level | 30-45 mL/min/1.73 m² | 1504 | 0.26 (-1.29 to 1.82) |
| | 45-60 mL/min/1.73 m² | 8476 | 1.69 (0.87 to 2.52) |
| | 60-90 mL/min/1.73 m² | 98566 | 1.20 (0.91 to 1.49) |
| | ≥ 90 mL/min/1.73 m² | 83606 | 0.87 (0.61 to 1.12) |
| Potassium level | < 3.5 mmol/L | 8953 | 0.83 (0.12 to 1.53) |
| | 3.5-4.0 mmol/L | 73443 | 1.18 (0.83 to 1.52) |
| | 4.0-4.5 mmol/L | 88652 | 1.04 (0.78 to 1.30) |
| | ≥ 4.5 mmol/L | 21102 | 0.92 (0.35 to 1.49) |

The subgroup-level excess risk predictions are based on the average individual-level risk prediction of the (full) 66-covariate model and calculated using augmented inverse propensity weighting.
